# Immune-developmental processes contribute to schizophrenia risk: insights from a genetic overlap study with height

**DOI:** 10.1101/2024.04.10.24305626

**Authors:** Cato Romero, Christiaan de Leeuw, Marijn Schipper, Bernardo de A.P.C. Maciel, Martijn P. van den Heuvel, Rachel M. Brouwer, August B. Smit, Frank Koopmans, Danielle Posthuma, Sophie van der Sluis

## Abstract

**Background:** Shorter stature has been phenotypically linked to increased prevalence of schizophrenia (SCZ), but the nature of this association is unknown.

**Methods:** Using genome-wide genetic data, we studied the SCZ-height relationship on a genetic level. Applying novel genetic methods and tools, we analyzed gene-sets, tissue-types, cell-types, local genetic correlation, conditional genetic analyses, and fine-mapping of effector-genes to scrutinize the SCZ-height relationship.

**Results:** We identified 142 genes statistically associated with both SCZ and height and found enrichment in immune response gene-sets. Genetic annotations implicated the pituitary and specifically mesenchymal stem cells for height and thyrotropic cells for SCZ. While the global SCZ-height genetic correlation was nonsignificant, 9 genomic regions showed robust local genetic correlations (7 negative, 6 in the MHC-region). The shared genetic signal for SCZ and height within the 6 MHC-regions was partially explained by mutual genetic overlap with white blood cell count, particularly lymphocytes. Fine-mapping prioritized 3 shared effector-genes (*GIGYF2*, *HLA-C*, and *LIN28B*) involved in immune response sensitivity and development of immune and pituitary cell-types.

**Conclusions:** Overall, our findings suggest an involvement during height-development of thyrotropic cells and immune response sensitivity contributing towards risk of SCZ.

## Introduction

Shorter stature has been linked to a small but consistent association with schizophrenia (SCZ) ^1–9^, with some studies showing a 15% increase in SCZ prevalence in males of below-average height^10^. While adult height shows consistent associations with SCZ prevalence, evidence for an association with birth length is inconsistent^4,11^, prompting further investigation into the nature of the SCZ-height relationship. The association between adult height and hormonal imbalance^12^, brain size^13^, infections^14^, nutrition^15^, and socioeconomic status^16^ alludes to a developmental risk component in the SCZ-height relationship, as factors influencing physical growth may interact with SCZ pathogenesis. Despite availability of large-scale genome-wide association studies (GWAS), few studies have investigated the genetic SCZ-height relationship directly. Current genetic evidence suggest certain SCZ-linked deletions (e.g., 22q11.2, 1q21.1, and 3q29) reduce height^17^. However, previous genome-wide association studies (GWAS) observed sign-discordant as well as sign-concordant shared SCZ-height signal^18^, and some SCZ-linked duplications (e.g., 16p11.2) associate with an *increase* in height^19^. These findings point to the existence of a complex genetic relationship between SCZ and height that can contribute to the observed negative phenotypic association. Leveraging larger GWAS datasets and novel methods, we aimed to further investigate the extent and nature of genetic overlap between SCZ and height, and to identify biological processes underlying this relationship (**Supplementary** Figure 1 displays the study design).

## Methods and Materials

### Genome-wide association study of height using UK Biobank

We conducted GWASs on height using the UK Biobank (UKB) data. Prior to analyses, we excluded variants with high missingness (>0.05), low imputation score (INFO value <0.9) and low minor allele frequency (MAF < 0.01), leaving 8,515,957 genetic variants. Subjects were filtered on genetic relatedness and European ancestry, resulting in an available sample size of 382,754 participants (54% female). Covariates were standardized and included age, sex, array and the first 20 principal components of population structure. This study was conducted under UKB application number 16406. Based on cohort descriptions, there is no sample overlap between the SCZ, UKB-height and GIANT-height samples.

### Global genetic correlations and SNP-based heritability estimation

We applied bivariate Linkage-Disequilibrium Score (LDSC) regression^20^ to estimate the genome-wide genetic correlation (*r_g_*) between SCZ and height, utilizing precomputed LD scores from the 1000 Genomes data (for each of the European, East-Asian, Latino, and African ancestries, separately) and filtering GWAS summary statistics to HapMap3 SNPs (**Data availability**). LDSC regression, requiring only GWAS summary statistics, provides *r_g_*-estimates unbiased by sample overlap.

### MiXeR estimation of shared genetic architecture beyond genetic correlations

To estimate shared genetic architecture, MiXeR^21^ analyzes GWAS summary datasets using a Gaussian mixture model, assuming a “mixture” of causal and non-causal variants that each follow a distinct Gaussian distribution. The model accounts for factors such as LD structure, genomic inflation from cryptic relatedness, MAF, GWAS sample size, and sample overlap. In this study, we used bivariate MiXeR to analyze SCZ and height in both full and sex-stratified samples. To prevent bias in polygenicity estimates caused by complex LD, the MHC region was excluded from all GWAS prior to analysis. All AIC and BIC fit indices were positive, indicating sufficient fit.

### Overlapping SNPs, genes, and gene-set enrichment analyses

To identify genome-wide significant (GWS) SNPs shared between SCZ and height, we filtered GWAS summary statistics of SCZ (7,193,791 SNPs), UKB-height (8,515,957 SNPs) and GIANT-height (2,550,858 SNPs) to 832,763 overlapping SNPs. SNPs meeting the GWS threshold (*P* < 5e-8) in SCZ and UKB-height were extracted, with semi-replication using GIANT-height (*α*_BON_ = 1.23 × 10^-4^). Fisher’s exact tests evaluated whether overlap exceeded chance levels. LD structure was addressed by clumping shared GWS SNPs using PLINK 1.9^22^, defining independent loci (LD *r*^2^ < 0.2, 250kb). MHC lead SNPs were further clumped using progressively larger (100kb) window sizes to account for long-range LD. Effect concordance was assessed through lead SNP beta coefficients. LD correlation structure was obtained through 10K UKB genotypes (**Data availability**).

SNP-to-gene annotation used Multi-marker Analysis of GenoMic Annotation (MAGMA)^23^, which calculated gene-based *P*-values accounting for LD between SNPs (19,427 genes; GRCh37). Shared genes were defined by Bonferroni-corrected significance (*α_BON_* =.05/19,427= 2.57 x 10^-6^). Functional enrichment of shared genes was analyzed in Functional Mapping and Annotation of Genome-Wide Association Studies (FUMA)^24^, testing 34,550 MSigDB^25^ v6.2 gene-sets with correction for multiple testing (*α_BON_* = .05/34,550 = 1.45 x 10^-6^). MAGMA also assessed gene-set and gene-property annotations, testing 50 hallmark gene-sets, 53 tissue-types (GTEx v8^26^), and 25 pituitary cell-types^27^ with combined Bonferroni correction (*α_BON_*=.05/128 = 3.90 × 10^-4^)

### Local genetic correlation analysis of SCZ and height

Local Analysis of [co]Variant Annotation (LAVA) identifies genomic regions with correlated genetic signals between traits^28^. LAVA requires GWAS summary statistics, estimated sample overlap (using bivariate LDSC intercepts), an external LD reference (1000 Genomes v3; **Data Availability**), and predefined genomic regions. Using LAVA’s default genomic region definitions, 2,517 genomic regions were defined for European data. LAVA’s workflow involves two steps: filtering regions with sufficient genetic signal in both traits (default local *h*^2^_SNP_; *P* < 1 x 10^-4^) and estimating bivariate local *r*_g_ in selected regions. Of the 2,517 regions, 816 showed sufficient genetic signal for both SCZ and height. Bivariate local *r*_g_ *P*-values were Bonferroni-corrected for multiple testing (*α_BON_* = 3.13 x 10^-5^; two-sided)

### Fine-mapping and gene prioritization in regions associated between SCZ and UKB-height

Fine-mapping of the 9 genomic regions that significantly correlated between SCZ and UKB-height was conducted using FINEMAP^29^, with LD structure estimated from 100K unrelated UKB subjects. Due to the complexity of fine-mapping the MHC region, FINEMAP was restricted to model 1 causal SNP per locus. To prioritize genes from FINEMAP’s 95% credible sets, we used FLAMES^30^ (Fine-mapped Locus Assessment Model of Effector geneS; v1.0.0), which predicts effector genes mediating trait associations based on locus-specific SNP-to-gene links and gene network convergence. Genes with FLAMES confidence scores > 0.05 were considered likely effector genes. Shared effector genes between SCZ and UKB-height were defined as those with FLAMES scores > 0.05 for both traits. We ran FLAMES using the default settings, i.e., only annotating SNPs to coding genes mapped within 750kb of the SNPs.

For *HLA-C*, we assessed expression and regulation in dorsolateral prefrontal cortex proteomics data from 47 SCZ cases versus 49 controls (Koopmans et al., under review, PRIDE dataset ID: PXD058441).

### Conditional local genetic correlation analysis in LAVA

We used conditional local genetic analysis in LAVA to assess whether the genetic overlap between SCZ and UKB-height in correlated regions was influenced by other covariates. This method tests whether the marginal genetic correlation (*r*_g_^marginal^) between two traits (*Y* and *X_1_*) significantly differs from the conditional estimate (*r*_g_^conditional^) after adjusting for a covariate (*X_2_* ). If *r_g_*^marginal^ ≠ *r_g_*^conditional^, the covariate significantly impacts the genetic covariance. Significance was determined by the conditional genetic covariance (*Cov*_g_; details in Supplementary Note 9.). Defined as a regression model, either SCZ or height must be chosen as the outcome (*Y*). Based on Mendelian Randomization SCZ was chosen as the outcome.

To be assessed, a covariate required significant local univariate genetic signal (*P* < 1 x 10^-4^) and genetic correlation with SCZ and height (*α_BON_* =.05/9 = 5.55 x 10^-3^). Applying these filters, 25 out of 44 covariates could be assessed. We tested 25 covariates across 9 loci, correcting for multiple testing (*α_BON_* = .05/(9 x 25) = 2.06 x 10^-4^). Covariates with significant effects were further replicated in SCZ and Giant-height analyses. Sample overlap estimates for LAVA were calculated using LDSC.

### Replication in non-European ancestries

We attempted to replicate our findings in East-Asian, African, and Latino ancestry GWAS^31,32^ data. First, we tested the overlap of 20 lead SNPs, 142 MAGMA genes, and gene-property results (pituitary/mesenchymal cells for height; pituitary/thyrotropic cells for SCZ), applying Bonferroni corrections (*α_BON_*=*α/nr. Lead SNPs* = .05/20 = 2.50 × 10^-3^; MAGMA genes: *α_BON_*=*α/nr. genes* = .05/142 = 3.52 × 10^-4^). Due to insufficient sample sizes in Latino (N_SCZcases_ = 1,234; N_height_ = 58,709) and African (N_SCZcases_ = 6,152; N_height_ = 168,193) GWASs, only East-Asian ancestry GWASs (N_SCZcases_ = 14,004; N_height_ = 363,856) were viable for local genetic correlation replication. To account for differences in LD structure, we used the LAVA partitioning algorithm on 1000 Genomes (v3) East-Asian LD reference. We focused on the 9 genomic regions significantly associated with SCZ and height in European analyses and identified 11 overlapping East-Asian regions for replication.

## Results

### Phenotypic association, Height GWAS and shared genetic architecture with SCZ

Using data from the UK Biobank (UKB), we confirmed the reported association between SCZ and shorter height in both sexes, although the lower genetic liability of SCZ cases in UKB limits the generalizability of our findings^33^ (**Supplementary Notes 1–2; Supplementary** Figures 2A–C**; Supplementary Table 1**). Next, we sought to explore how common genetic variation accounts for this observed phenotypic relationship. We obtained SCZ GWAS summary statistics of European (EUR) ancestry individuals from the Psychiatric Genetics Consortium^31^ (N_eff_^34^ = (4 x N_cases_ x N_controls_)/N_total_ = 157,066). We conducted a GWAS on height using EUR-ancestry data in the UKB (UKB-height; **Methods**). Although larger height GWAS exist, their sparse SNP coverage (∼1 million SNPs) adversely affects genetic overlap analyses (e.g., a substantial proportion of LAVA loci would fail). The UKB data, with its denser coverage (∼8 million SNPs), ensures more robust and comprehensive analyses of genetic overlap with SCZ. (**Supplementary Table 2** displays all SCZ and height GWAS data used in this study). Using LDSC regression^20^ (**Methods**), we estimated the genome-wide genetic correlation between SCZ and UKB-height to be near-zero and non-significant (*r*_g_ = -.01, SE = .02, *P* = .72), which is in line with previous reports^18^. MiXeR^21^, unlike LDSC, quantifies shared genetic architectures between traits beyond the global genetic correlation. Bivariate MiXeR (**Methods**) estimated ∼1,100 shared SNPs (SE = 200) contributing to shared genetic architecture between SCZ and UKB-height (12% and 35% of all SCZ and UKB-height MiXeR estimated SNPs, respectively; **Figure 1A**-**D**). The sign-concordance of shared SNPs was 48% (SD = 1.0%), consistent with a near-zero global genetic correlation.

**Figure 1.**
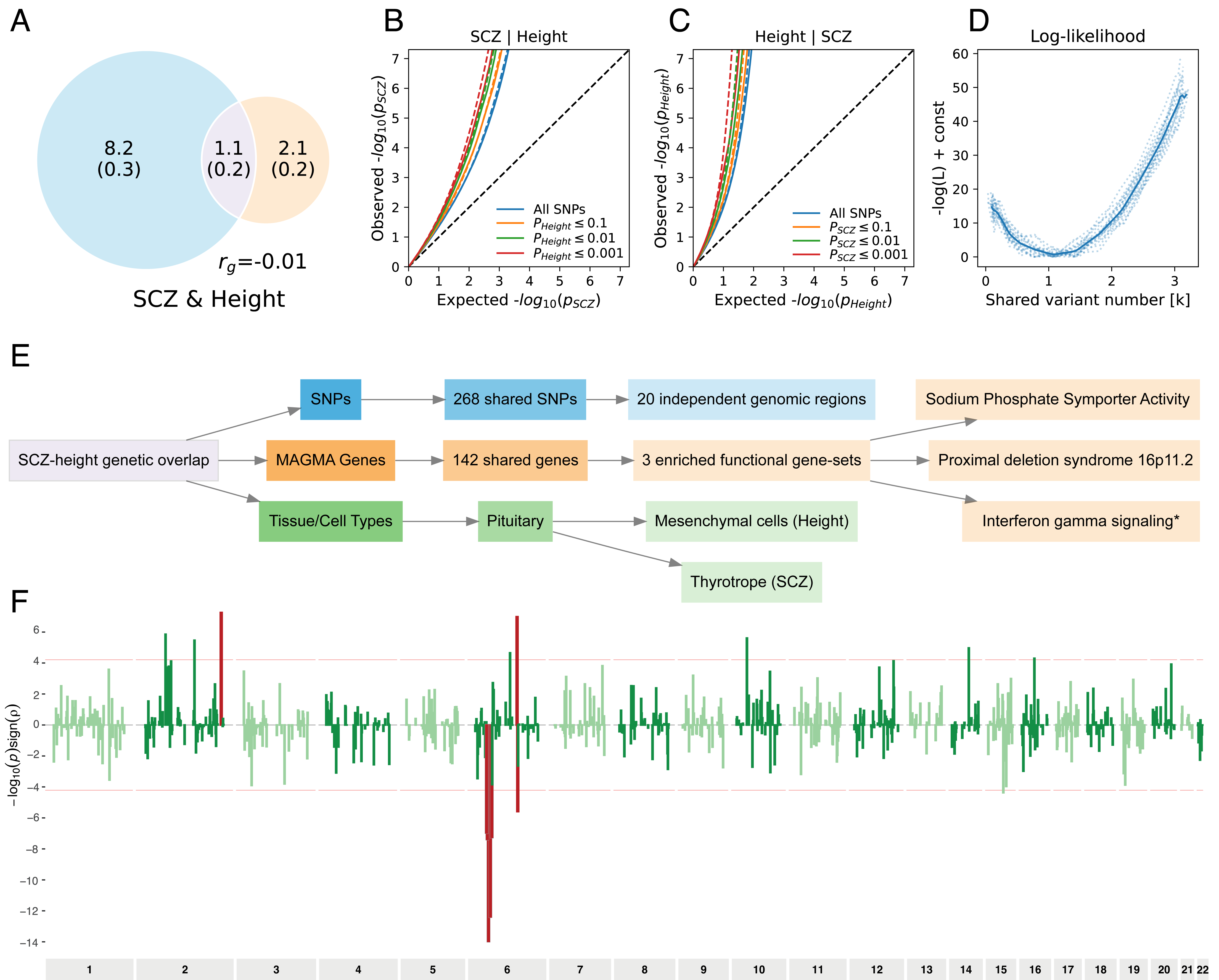
Shared genetic overlap between SCZ and height. 1A) Venn-diagram of estimated MiXeR overlap of shared SNPs between SCZ and UKB-height. Jaccard index (ratio of overlap to total area) is 9.65%. Estimates are in 1000 SNPs, i.e., 1.1 refers to 1,100 shared SNPs with SE = 200. B-C) Conditional Q-Q plot of expected versus observed -log10(P) compares the distribution of p-values for SNP associations with SCZ | UKB-height SNPs (B) and UKB-height | SCZ SNPs (C) against the uniform null expectation (x-axis). The diagonal dashed line represents the null hypothesis, where deviation by observed p-values represent significant association for SNPs with P < 0.001, 0.01, 0.1, and 1, respectively. D) Negative log-likelihood plot as function of polygenic overlap. Lowest point of line indicates number of shared variants with optimal fit. E) Flowchart summarizing genetic overlap between SCZ and height regarding specific SNPs, independent risk loci, shared MAGMA genes, gene-set enrichment, tissue and cell type results. Gene-sets with * are not significant after exclusion of the MHC region F) Local genetic correlations between SCZ and UKB-height over all loci in the genome. Y-axis is defined as -log10(P)*sign of rho. Red lines represent local genetic correlations that replicated between SCZ and GIANT-height.

### Genetic variants and functional annotations shared between SCZ and height

We investigated specific GWS SNPs, genes and biological annotations shared between SCZ and UKB-height and then used an independent height dataset (GIANT-height^35^; N_GIANT-height_ = 253,288) for semi-replication. That is, for the replication we used the same SCZ GWAS summary statistics (as such data are not available for a sample of similar size and ancestry) but different height data. Analyses were conducted with and without the MHC region (chromosome 6 28,477,797 – 33,448,354 in hg19) to address possible inflation in overlap due to high gene-density and structural complexity. Among the GWS SNPs from SCZ (2,469; 176 independent risk loci) and UKB-height (23,567; 1,226 independent risk loci), we observed 408 mutually associated SNPs, out of which 268 SNPs were semi-replicated and sign-concordant (*α_BON_*=.05/408 = 1.23 × 10^-4^) in GIANT-height, which is above the expected chance level (OR = 3.68, Fisher’s exact test *P* = 1.18 x 10^-32^). Clumping yielded 20 independent risk loci (5 within the MHC), and 55% of their lead SNPs showed concordant effects (**Supplementary Table 3**). Next, we performed genome-wide gene-based association analysis using MAGMA (**Methods**)^23^. We identified 558 GWS genes for SCZ and 2,883 for UKB-height. Of these, 205 overlapped, and 142 replicated in GIANT-height (*α_BON_*=.05/205 = 2.44 × 10^-4^), which is again significantly above chance level (OR = 2.01, Fisher’s exact test *P*-value = 2.87 × 10^-11^, without MHC genes OR = 1.89, Fisher’s exact test *P* = 5.33 × 10^-8^, **Supplementary Table 4**). Significant SNP and/or gene-overlap was also seen between height and other psychiatric disorders. Specifically, SNP and gene-overlap with height was significant for bipolar disorder^36^ (BIP; N_eff_ = 49,366), SNP-overlap was significant for insomnia disorder^37^ (N_eff_ = 313,722) and autism spectrum disorder^38^ (N_eff_ = 44,393), but overlap was not significant for attention deficit/hyperactivity disorder^39^ (N_eff_ = 48,990), alcohol use disorder^40^ (N_eff_ = 145,999), major depressive disorder^41^ (N_eff_ = 156,547), and anorexia nervosa^42^ (N_eff_ = 52,027; **Supplementary Table 5**). Out of the genes shared between BIP and height, 90% (N = 18) were also shared between SCZ and height, suggesting concordance in SCZ-BIP overlap with height.

Convergence of the 142 genes that were associated to both SCZ and height onto gene-sets was investigated using GENE2FUNC in FUMA (**Methods**)^24^. Gene-sets were obtained from MSigDB^25^ (N_MSigDB gene-sets_ = 34,550). Significant enrichment was found for 7 gene-sets (*α_BON_* =.05/34,550 = 1.45 x 10^-6^; **Figure 1E**), including 4 positional gene-sets (*chr16p11, chr3p21, chr6p21*, and *chr6p22*), and 3 functional gene-sets (*sodium phosphate symporter activity, interferon gamma signaling* and *proximal deletion syndrome*; although *chr6p21* and *interferon gamma signaling* were non-significant when MHC genes were excluded; **Supplementary Table 6**). Gene-set enrichment analysis of mutual GWS genes is an incomplete assessment of functional convergence as different genes can partake in the same biological processes or be expressed in the same biological tissue. To investigate the existence of additionally significant gene-sets, as well as tissue enrichment, we conducted gene-set and gene-property analysis in MAGMA on both SCZ and height (using the biologically-established and non-redundant hallmark gene-sets N_hallmark gene-sets_ = 50 from MsigDB v6.2^25,43^ and N_tissue-types_ = 53 from GTEx v8.0^26^) with the entire set of 19,427 MAGMA genes. While no additional gene-sets were significant, gene-property analysis of tissue-types found *the pituitary* as significantly associated with both SCZ and UKB-height. Follow-up cell-type analysis within the pituitary (N_cell-types_ = 25 from Zhang et al.^27^) found significant enrichment for *mesenchymal stem cells* (MSC) for UKB- and GIANT-height, and *thyrotrophic cells* (TC) for SCZ. All gene-property results were significant after correcting for multiple testing and after excluding the MHC region (*α_BON_* =.05/(50 + 53 + 25) = 3.90 × 10^-4^; **Supplementary Table 7**). In summary, multiple risk loci and genes are associated with both phenotypes, implicating shared biological processes between these traits.

### Investigating genetic correlations within genomic regions

Shared genetic architecture with concordant and discordant SNP effects suggest the possibility of significantly shared *local* or regional genetic signal. To evaluate shared local signal, we conducted local genetic correlation analysis using LAVA (**Methods**)^28^. To filter out clearly non-associated loci, only genomic regions with suggestive (LAVA default *P* < 1 x 10^-4^) univariate genetic signal in both SCZ and height were considered for estimation of local bivariate genetic correlations, resulting in 816 out of 2,517 predefined, roughly independent genomic regions. Subsequently, bivariate genetic correlation was evaluated in these 816 regions and 16 regions showed significant SCZ-height local correlation after correcting for multiple testing (*α_BON_* = .05/816 = 6.13 x 10^-5^; **Table 1**; **Figure 1F**). Semi-replication with the independent GIANT-height data evinced significant associations in 9 of these 16 regions (*α_BON_* = .05/16 = 3.13 x 10^-3^; **Supplementary** Figure 3A-B). Among these 9 semi-replicated significant regions, 2 were positively and 7 were negatively correlated. Of these 7, 6 were localized within the MHC region on chromosome 6 (all but two regions (chromosome 6 104,945,107 – 106,052,133 and chromosome 6 107,319,938 – 109,273,876) overlap with the aforementioned 20 shared lead SNPs).

**Table 1.** Genomic regions showing significant local genetic correlation between SCZ and height. Note: All semi-replicated results showed sign concordance with the main analysis. (*) indicates region within the MHC region on chromosome 6.

| Locus number | CHR | Start | Stop | Rho | P-value | Semi-replication P-value |
| --- | --- | --- | --- | --- | --- | --- |
| 410 | 2 | 233360693 | 234089948 | 0.56 | $5.50 \times 10^{-8}$ | $3.57 \times 10^{-5}$ |
| 970(*) | 6 | 30316328 | 30932188 | -0.45 | $8.74 \times 10^{-5}$ | $1.34 \times 10^{-5}$ |
| 971(*) | 6 | 30932189 | 31143244 | -0.46 | $4.08 \times 10^{-8}$ | $8.20 \times 10^{-7}$ |
| 972(*) | 6 | 31143245 | 31250172 | -0.66 | $1.06 \times 10^{-14}$ | $8.13 \times 10^{-6}$ |
| 973(*) | 6 | 31250173 | 31320268 | -0.64 | $2.12 \times 10^{-12}$ | $3.61 \times 10^{-5}$ |
| 974(*) | 6 | 31320269 | 31577220 | -0.64 | $4.04 \times 10^{-13}$ | $1.66 \times 10^{-6}$ |
| 981(*) | 6 | 32636612 | 32682213 | -0.51 | $5.53 \times 10^{-8}$ | $3.93 \times 10^{-5}$ |
| 1050 | 6 | 104945107 | 106052133 | 0.64 | $1.03 \times 10^{-7}$ | $6.13 \times 10^{-5}$ |
| 1052 | 6 | 107319938 | 109273876 | -0.41 | $2.58 \times 10^{-6}$ | $7.00 \times 10^{-7}$ |

We sought to investigate multiple additional aspects of the local SCZ-height genetic relationship, including the extent of sex-specific effects, non-linear SCZ-height associations across height strata, and evidence against indirect genetic effects (e.g., assortative mating, population stratification, and gene-environment interactions). In summary, we detected 6 additional local genetic correlations between SCZ and height specific to males using UKB-height data, of which only 1 semi-replicated (chromosome 15 89,385,687 – 90,632,694) in male-stratified GIANT-height data^44^. No female-specific local genetic correlations were observed, and concordant effects between sexes were confirmed using MiXeR on sex-stratified SCZ-height overlap (**Supplementary Note 3; Supplementary Table 8**). Within the 9 genomic regions in which we observed local genetic correlations, the association between SCZ and height appeared to be linear, except for one region (chromosome 6 107,319,938 – 109,273,876) where the effect was primarily driven by the 20% shortest individuals (**Supplementary Note 4; Supplementary Table 9**). Using a within-sibling GWAS on height^45^, which can better control for indirect genetic effects, 4 of the 9 robust regions were semi-replicated, lending evidence to associations being based on direct (rather than indirect) genetic effects in these regions (**Supplementary Note 5**; **Supplementary Table 10**).

Next, we used FLAMES (**Methods**)^30^ to prioritize which genes most likely underlie the local genetic SCZ-height correlations in the 9 robust regions: three genes — *GIGYF2*, *HLA-C* and *LIN28B* — converged between SCZ and UKB-height as the most likely genes underlying the genetic correlation in three genomic regions (chromosome 2 233,360,693 – 234,089,948, chromosome 6 31,250,173 – 31,320,268, and chromosome 6 104,945,107 – 106,052,133, respectively; **Table 2, Supplementary** Figure 4; **Supplementary Note 6; Supplementary Table 11**). Gene-prioritization of MHC in SCZ has previously not supported a role for *HLA-C*^46^. However, look-up of *HLA-C* in SCZ differentially expressed proteins in dorsal lateral prefrontal cortex (dlPFC) found *HLA-C* to be significantly differentially regulated (log^2^(FC) = -0.89, *Q*-value = 0.02 (5% FDR, 5,242 proteins tested); Koopmans et al., under review; **Methods**) in 47 SCZ cases as compared to 49 controls. In the remaining 6 regions no or different genes were prioritized between SCZ and height.

**Table 2.** Top prioritized SNPs within SCZ-height correlated LAVA loci and allele frequencies. Note: Flames score above 0.05 is regarded as sufficient. MAF for multiple ancestries are based on gnomADv2.1.1. Which allele is the minor allele is based on the European ancestry reference panel, MAF values with * indicate that this allele is the major allele in the respective population, not minor allele. PIP = Posterior Inclusion Probability; nSNP in set refers to the number of SNPs in the 95% credible set, sQTL: splicing quantitative trait locus, eQTL: expression quantitative trait locus, HI-C: chromatin conformation capture, DHS-promoter: DN-ase I hypersensitive site-promoter.

| Locus | Trait | Flames Gene | Flames Score | Top SNP | PIP | nSNP in set | SNP on Gene | EUR MAF | EAS MAF | AFR MAF | LAT MAF |
| --- | --- | --- | --- | --- | --- | --- | --- | --- | --- | --- | --- |
| 410 | Height | *GIGYF2* | .05 | rs283477 | .12 | 75 | sQTL, eQTL, intron variant | .31 | .36 | .29 | .30 |
|  | SCZ | *GIGYF2* | .11 | rs778371 | .16 | 46 | sQTL | .30 | .11 | .24 | .31 |
| 970(*) | Height | *SFTA2* | .21 | rs2253718 | .93 | 6 | eQTL | .36 | .07 | .41 | .20 |
|  | SCZ | *DDX39B* | .12 | rs1264347 | .10 | 69 |  | .12 | .01 | .04 | .03 |
| 971(*) | Height | *MUC22* | .24 | rs9262549 | .99 | 1 | Missense variant | .30 | .28 | .12 | .21 |
|  | SCZ | *TRIM39* | .04 | rs1634726 | .36 | 11 |  | .12 | .00 | .02 | .03 |
| 972(*) | Height | *HLA-C* | .16 | rs2524080 | .74 | 5 | sQTL, eQTL | .29 | .19 | .16 | .22 |
|  | SCZ | *HCG27* | .04 | rs1793894 | .73 | 3 | eQTL | .13 | .003 | .09 | .06 |
| 973(*) | Height | *HLA-C* | .06 | rs2524054 | .45 | 5 | sQTL, eQTL | .27 | .19 | .09 | .17 |
|  | SCZ | *HLA-C* | .10 | rs2923008 | .13 | 31 | sQTL, eQTL | .12 | .004 | .02 | .04 |
| 974(*) | Height | *HLA-B* | .02 | rs2523578 | .36 | 3 | sQTL, eQTL | .26 | .03 | .20 | .13 |
|  | SCZ | *ATP6V1G2* | .03 | rs2596495 | .11 | 15 | DHS-promoter | .13 | .004 | .11 | .06 |
| 981(*) | Height | *HLA-DQB1* | .04 | rs9275109 | .50 | 5 | sQTL, eQTL, Hi-C | .37 | .15 | .36 | .20 |
|  | SCZ | *HLA-DQA1* | .03 | rs1617322 | .10 | 15 | sQTL, eQTL | .13 | .06 | .08 | .06 |
| 1050 | Height | *LIN28B* | .34 | rs314265 | .17 | 11 | eQTL | .23 | .23 | .41 | .25 |
|  | SCZ | *LIN28B* | .14 | rs457286 | .07 | 52 | eQTL, intron variant | .44 | .33 | .77* | .33 |
| 1052 | Height | *FOXO3* | .24 | rs9400239 | .16 | 20 | Intron variant | .32 | .30 | .77* | .40 |
|  | SCZ | *SNX3* | .05 | rs11964401 | .09 | 42 |  | .49 | .36 | .83* | .49 |

### Assessing involvement of external traits on correlated genomic regions

We assessed whether the 9 local genetic SCZ-height correlations could be further understood through shared genetic covariance with other traits, that is, whether the genetic relationship between SCZ and height is mediated by other traits. To this end, we conducted conditional local genetic correlation analyses in LAVA (**Methods; Supplementary Note 7**). GWAS summary statistics of potential covariates were selected based on their known epidemiological relation to SCZ or height to: 1) investigate whether the genetic SCZ-height overlap is shared with related traits (e.g., BIP and body-mass index), 2) assess involvement of potentially confounding factors (e.g., social deprivation), and 3) assess potential biological implications in the relationship between SCZ and height (e.g., (sex-)hormones, and metabolites; full list of all 44 selected covariates and their sources in **Supplementary Table 12**). Subsequently, covariates were filtered for having significant univariate genetic signal (LAVA default *P* < 1 x 10^-4^) and significant genetic correlation with both SCZ and UKB-height in at least 1 of the 9 genomic regions (*α_BON_* = 0.05/9 = 5.55 x 10^-3^). After filtering, 25 covariates remained.

Specified as a mediation model, conditional genetic analyses require the appointment of a predictor and an outcome variable. Generalized Summary-data-based Mendelian Randomization (GSMR)^47^ uses genetic information to determine, for genetically and phenotypically correlated traits, the most likely direction of causation. Here, we used GSMR to determine whether SCZ or UKB-height should be set as outcome variable in our local genetic mediation analyses. GSMR yielded stronger evidence, both for p-value and standardized effect size, for UKB-height predicting SCZ (B_height_ = -.024, SE = .006, P = 2.00 x 10^-5^) than for SCZ predicting UKB-height (B_SCZ_ = -.020, SE = .008, P = .02), hence in our mediation analyses, the effects of the 25 covariates were assessed with SCZ as the outcome variable and UKB-height as predictor (**Supplementary Note 8**). Among related or confounding traits, only conditioning on BIP resulted in a large (sub-threshold) significant decrease in the genetic correlation between SCZ and UKB-height within 4 loci (**Figure 2**). Among traits potentially sharing biological processes with both SCZ and height, conditioning on the genetic signal of white blood cell subtype counts (lymphocyte and neutrophil counts), significantly decreased the genetic correlation between SCZ and UKB-height in 6 out of 9 genomic regions, all residing within the MHC region. Lastly, conditioning on the genetic effects of intra-cranial volume strongly affected the SCZ-height relation on chromosome 6 outside MHC. All the effects of covariates remained significant in semi-replication using GIANT-height GWAS summary statistics (**Supplementary Table 13; Supplementary** Figure 5-6 shows results from model with height as outcome).

**Figure 2.**
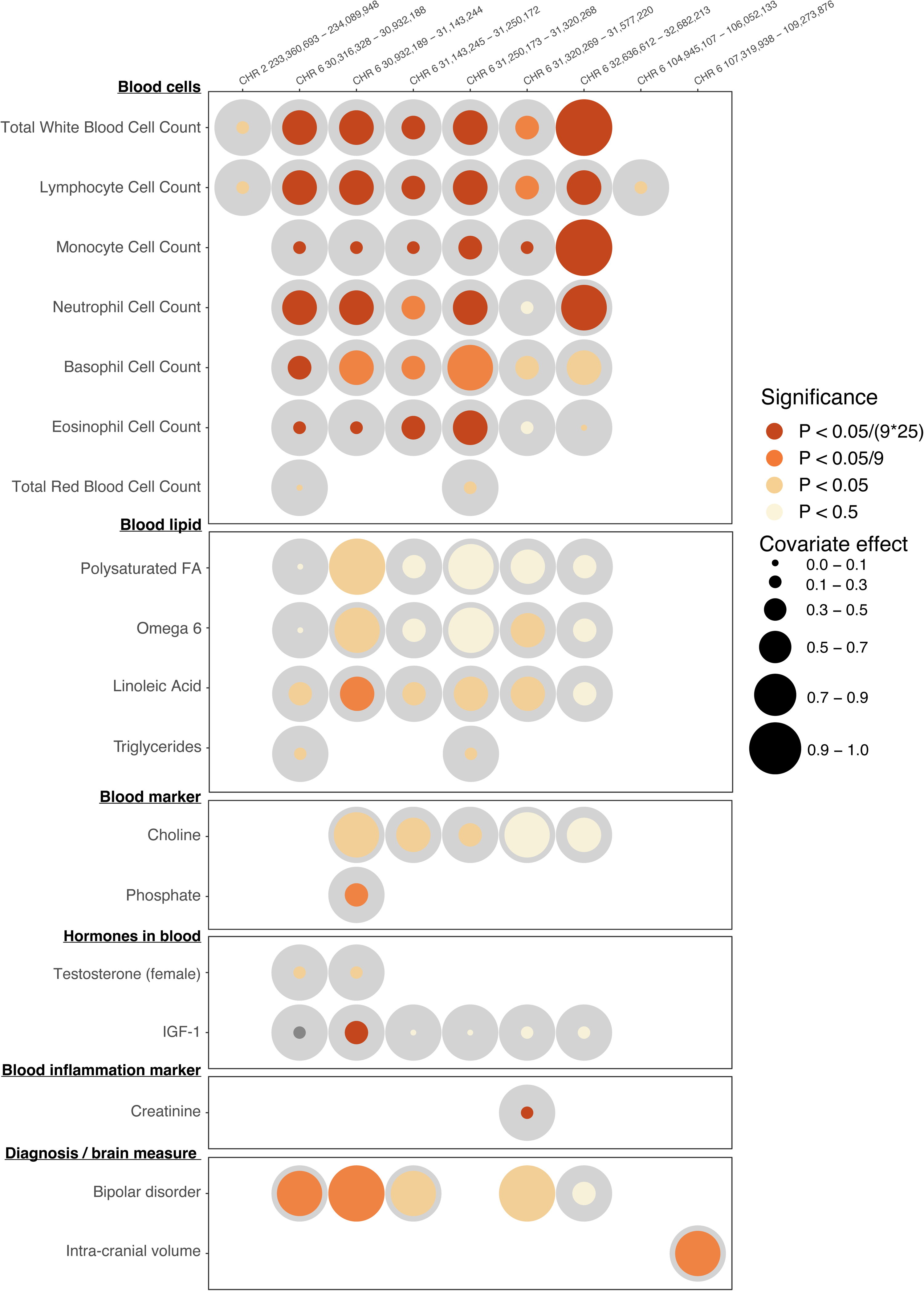
Results from conditional local genetic correlation analyses. Results of the conditional local genetic correlation analyses for covariates that showed (nominal) significant conditional effects. Left side of the plot shows covariate name and category; top side gives genomic region definitions. Circle sizes represent the estimated covariate effect. The covariate effect is the genetic covariance between the covariate trait on the one hand and the covariance shared between SCZ-height on the other. Grey circles represent theoretical complete overlap between covariate and SCZ-height, while colored circles represent the observed covariate – SCZ-height overlap within each of the 9 genomic regions. To illustrate, for Total white blood cell count in region chromosome 6 32,636,612 – 32,682,213 (top panel, rightmost circle), all genetic covariance shared between SCZ and height is also shared with white blood cell count, while in the other regions the genetic signal is only partly, yet often significantly, shared. Colors indicate significance of the covariate trait estimate: light yellow = not significant, light orange = nominally significant, orange = significant corrected for number of genomic regions tested (9), and dark red = significant corrected for number of genomic regions and number of covariates tested (9 x 25). This figure uses absolute covariate effect, for the sign of the covariate effect, please see **Supplementary table 13**.

To follow-up the genetic relation between SCZ, height and white blood cell count, we conducted analyses in UKB data to assess whether phenotypic relationships mirror the observed genetic relations. Results indeed showed a negative phenotypic relation of white blood cell count (in 10^9^ cells/liter) with UKB-height (in cm; β = -.13, SE_β_ = 2.01 x 10^-3^, B = -.03, SE_B_ = 4.66 x 10^-4^, T = -63.71, P < 2.00 x 10^-16^), and a positive relation with SCZ diagnosis (β = .02, SE_β_ = 1.46 x 10^-3^, B = .82, SE_B_ = .06, T = 14.39, P < 2.00 x 10^-16^), mirroring previous reports^48,49^.

### Replication in non-European ancestry

Lastly, to evaluate whether the genetic overlap between SCZ and height observed in EUR-ancestry is evident across other ancestries, we analyzed GWAS data for East-Asian (EAS), African (AFR), and Latino (LAT) ancestry. Global genetic correlations were non-significant for EAS and LAT but positive for AFR (*r*_g_ = .29, SE = .13, P = .02). Replication of EUR-ancestry lead SNPs, genes, and annotations showed limited significance in EAS data and none in AFR or LAT data (see **Supplementary Note 9; Supplementary Tables 3, 4, 7**). Using EAS-based summary statistics for SCZ (N_eff_ = 30,523) and height (N = 363,856), none of the nine EUR-significant genomic regions replicated (**Supplementary Note 10; Supplementary Table 14**). While small SCZ samples affect the power of these cross-ancestry replication attempts, top SCZ EUR prioritized SNPs did show lower MAF in EAS ancestry (**Table 2, Supplementary Table 15**) suggesting potential ancestry-specific genetics effects.

## Discussion

We investigated the SCZ-height genetic relationship via multiple post-GWAS methods, replicating 6 of 8 previously reported shared lead SNPs^18^, and observe 14 addition lead SNPs, 142 shared genes, and several shared biological properties. Our findings paint a complex genetic picture, related to immune response and pituitary-driven development. The shared gene-set *sodium phosphate symporter activity* is involved in processes like nutrient absorption, cellular signaling, and skeletal development^50^. Although SCZ and height were enriched in different cell-types in the pituitary, these cell types are known to interact: thyroid stimulating hormone secreted by pituitary thyrotropic cells regulates mesenchymal cell differentiation^51^. Shared prioritized genes, *LIN28B*, *GIGYF2*, and *HLA-C*, appear to modulate inflammation and thyroid function, potentially explaining how SCZ’s dysregulated thyroid hormones can impact growth. Collectively, findings align with *the developmental risk factor model of psychosis*, suggesting that disruptions in growth- and immune-related processes may heighten susceptibility to SCZ^52^. For full discussion, see **Supplementary Note 11**.

Our data indicate that SCZ-height genetics may be more pronounced than in other psychiatric disorders. Gene and SNP overlap was observed for BIP, and BIP showed significant conditional effects in 4 loci, but most of this overlap was shared with SCZ, highlighting the strong genetic correlation (.67) between BIP and SCZ^53^ rather than BIP-specific components, an interpretation substantiated by non-significant associations between BIP and height in epidemiological literature^54^. However, statistical power needs to increase for other psychiatric disorders before we can verify the specificity of SCZ’s genetic association to height.

The aforementioned biological processes represent a plausible hypothesis concerning the relationship between SCZ and height; however, caution is needed regarding the generalizability of this hypothesis. While our results suggest that the hypothesis generalizes to both sexes, results did not replicate in non-EUR ancestries. To our knowledge, no epidemiological study has been published on the SCZ-height association in non-EUR samples, and our non-EUR UKB sample lacked statistical power to detect a mean height difference (**Supplementary Note 9**). Replication is hampered by power issues relating to smaller non-EUR (especially SCZ) GWAS samples, and lower MAF of the EUR lead SNPs in non-EUR ancestries (**Table 2; Supplementary Table 15**). While variation in MAF between ancestries could suggest that the SCZ-height relationship is specific to, or more pronounced in, people of EUR ancestry, additional epidemiological and genetic research in non-EUR ancestry is required to test this hypothesis.

The MHC region, frequently appearing in our results, poses methodological challenges for GWAS studies. While we replicated results without the MHC where possible, LAVA and FLAMES results are affected by the strong LD in this region: long-range LD can cause interdependence among nonadjacent loci that is not accounted for in LAVA, rendering the independence of the 6 MHC loci uncertain. Additionally, while we used FINEMAP to prioritize SNPs in LAVA-defined regions, specialized methods (SNP2HLA and HLA*SNP) are better suited for fine-mapping the MHC but require individual-level data. Nevertheless, our prioritization of *HLA-C* for SCZ is supported by significant downregulation in dlPFC SCZ proteomic data, suggesting *HLA-C* may contribute to the MHC-SCZ association in addition to other, previously implicated genes.

In summary, despite a near-zero global genetic correlation, shared genetic signals between SCZ and height is evident. By leveraging the genetic overlap with height, our findings converge on the involvement of thyrotropic cells and immune response sensitivity in SCZ and highlights development-specific biological processes contributing towards the etiology of SCZ.

## Acknowledgements

C.R., B.A.P.C.M., M.P.vd.H., M.S., R.M.B., A.B.S., D.P. and S.v.d.S were funded by NWO Gravitation: BRAINSCAPES: A Roadmap from Neurogenetics to Neurobiology (grant no. 024.004.012 [to D.P.]). F.K. was supported by the Simons Foundation, SFARI Award 882976. D.P. and C.d.L. were funded by a European Research Council advanced grant (no. ERC-2018-AdG GWAS2FUNC 834057 [to D.P.]). M.P.v.d.H. is supported by NWO VIDI Grant 452-16-015 and the ERC Consolidator of the European Research Council Grant 101001062.

The analyses were carried out on the Genetic Cluster Computer, which is financed by the Netherlands Scientific Organization (NWO: 480-05-003), by the VU University, Amsterdam, the Netherlands, and by the Dutch Brain Foundation, and is hosted by the Dutch National Computing and Networking Services SurfSARA. This research has been conducted using the UK Biobank resource under application number 16406. We thank the numerous participants, researchers, and staff from many studies who collected and contributed to the data. We particularly express our gratitude to all UKB participants that have been so generous to share their data for analysis.

**Supplementary Note Figure 6** and **Supplementary** Figure 1 was created with BioRender.com with permission to publish.

## Author Contributions

C.R.: Conceptualization, formal analyses, methodology, visualization, writing. C.d.L.: Feedback, formal analysis, methodology. M.S.: Feedback, formal analysis. B.A.P.C.M.: Feedback, visualization. M.P.vd.H.: Feedback, resources. R.M.B.: Feedback, resources. A.B.S.: Resources. F.K.: Resources. D.P.: Feedback, funding acquisition, resources. S.vd.S.: Conceptualization, methodology, project administration, writing, supervision.

## Data availability

GWAS summary statistics were downloaded from link for SCZ and link for GIANT-height.

All GWAS summary statistics are based on Human Genome Build 37 (GRCh37/hg19).

Precomputed LD scores and HapMap 3 reference file were obtained from: <u>Link</u>

1000 Genomes reference data: Link

LD correlation structure was obtained through the LD information available from 10K UKB genotypes: https://www.uk10k.org

Gene information was obtained from the GeneCard database (v5.12.0 Build 702; https://www.genecards.org).

## Software Availability

FLAMES(v1.0.0) https://github.com/Marijn-Schipper/FLAMES

MAGMA (v.1.10) gene-based and gene-property analysis: https://ctg.cncr.nl/software/magma

LAVA(v0.1.0) local genetic correlations: https://ctg.cncr.nl/software/lava

PLINK (v1.9) https://www.cog-genomics.org/plink/

G*power**: <u>Link</u>**

