## Supplementary material for "Immune-developmental processes contribute to schizophrenia risk: insights from a genetic overlap study with height": scz-height.Suppl.docx

**Supplementary Note 1: Phenotypic schizophrenia-height association and power calculations**

Using data from the UK Biobank (UKB)^1^, we first sought to replicate the on average 1 centimeter difference in height between SCZ-cases and controls, as observed in Swedish males by Zammit et al^2^. We further conducted sex-stratified analyses to assess whether a similar difference was present in females. Standing height was defined by Data-field 50, and SCZ was defined as an ICD-10 diagnosis within F20.0-F20.9, identifying 1,360 SCZ cases and 498,495 controls. After excluding subjects of non-European ancestry (N = 24,666) or without ancestry data (N = 15,899), 1,105 SCZ cases (81.2%) and 459,545 controls (94%) remained. Due to insufficient statistical power (range .13-.45 using a two-sample t-test and alpha level of 0.05) in non-European ancestry groups, mean height differences for African, East-Asian, South-Asian, and Admixed-American ancestries were not tested but are reported in **Supplementary Note 3**. Outlying height values (1^st^/3^rd^ quartile ± 1.5 x IQR) were excluded by sex and SCZ status, removing 3.105 individuals (0.6%). Final samples included 1,096 SCZ cases (685 males, 411 females) and 455,089 controls (207,581 males, 247,508 females). Age-adjusted mean height differences between SCZ cases and controls were analyzed separately for males, females, and the (age- and sex-corrected) combined sample. Statistical power to detect a 1 cm difference was calculated using G*Power. Based on Zammit et al^2^, SCZ cases were expected to be 1 centimeter shorter than controls. To calculate the statistical power for the female-only, male-only, and full samples, we took the mean and standard deviation from the age-adjusted height values from the control group of the female-only, male-only, and full samples. Further, we estimated the power to detect a 1 centimeter decrease in height assuming the same SD in cases as seen in controls (see **Supplementary Note Table 1** below),

#### **Supplementary Note Table 1**: Descriptive information for the sex-stratified phenotypic SCZ-height analyses

|  | Mean_controls_ | SD_controls_ | N_controls_ | N_cases_ | Estimated power  α=.05 / α=.0167 |
| --- | --- | --- | --- | --- | --- |
| Female-only | 162.61 | 5.97 | 247,508 | 411 | .92/.84 |
| Male-only | 175.79 | 6.41 | 207,581 | 685 | .98/.95 |
| Full sample | 169.20 | 6.17 | 455,089 | 1,096 | .99/.99 |

*Notes: Means and SDs are age-adjusted and, in the case of the full sample, also sex-adjusted. Estimated power is reported for α=.05 and for the Bonferroni corrected α=.05/3=.0167.*

Power was calculated for a two-tailed test and α=.05 and for the Bonferroni-corrected α=.05/3=.0167, given the available sample sizes from the SCZ cases and controls. These power analyses resulted in an estimated statistical power of 92% (84%), 98% (95%), and 99% (99%) for the female-only, male-only and full sample for α=.05 and α=.0167, respectively.

After data preprocessing and power analysis, 1,096 SCZ-cases (62.5% males) and 455,089 controls (45.7% males) with European (EUR) ancestry remained. We replicated the expected height difference between SCZ-cases and controls in both females (.65cm difference, Cohen’s d = .11, *P* = 2.70 x 10^-2^; **Supplementary Figure 2A**) and males (1.30cm difference, Cohen’s d = .20, *P* = 1.26 x 10^-7^; **Supplementary Figure 2B**), as well as in the full sample (1.06cm difference, Cohen’s d = .17, *P* = 1.38 x 10^-8^; **Supplementary Figure 2C**; **Supplementary Table 1**). These significant phenotypic associations suggest that SCZ is related to relative height (i.e., expected height given relevant factors like sex) rather than absolute height (**Supplementary Note 2**).

After the linear regression analyses had been conducted, the realized statistical power was calculated using the observed mean difference in height rather than the assumed 1-centimeter difference, and this resulted in an observed statistical power of 93% (85%), 99% (98%), and 99% (99%) for the female-only, male-only, and full analyses for α=.05 and α=.0167, respectively. For the descriptive adjusted and unadjusted height values from each subgroup, see **Supplementary Table 1**.

### **Supplementary Note 2: Is relationship with SCZ based on absolute or relative height?**

How downstream analyses are performed depends on whether the relationship of SCZ with height is based on *absolute* height (the exact measure in centimeters) or *relative* height (a person’s height relative to external factors like sex and age).

Direct assessment of whether absolute or relative height associates with SCZ would preferably require phenotypic data on height and schizophrenia status in addition to including data on age and sex and preferably more demographic variables related to height (such as ethnicity and socioeconomic status). We were not able to access data of this kind, however, we can still assess the likelihood of the association being with absolute or relative height based on theoretical predictions. According to the study by Zammit et al.^2^, which was conducted in males only, every centimeter increase in height is associated with a 0.99 hazard ratio for developing SCZ, meaning that for every 1 centimeter decrease in height a person’s risk for developing SCZ increases with 1%. Females are on average ~13 centimeter shorter than males^3^ meaning that if SCZ is associated with *absolute* height, females would on average have a 13% increased risk of developing SCZ compared to males. Yet, this is inconsistent with the observation that SCZ is slightly less prevalent in females than in males (ratio 1:1.4 for females and males, respectively)^4^. Additionally, people with a height of 145 – 160 cm would be in a high risk (15-30% increased risk) group and the ratio of females to males in this group (at least within the UK Biobank) is approximately 15:1 (**Supplementary Note Figure 1A**).

If we assume relative height to be the correct measure, then height would have to be normalized within males and females before data are jointly analyzed, to make their height measures relative to their sex. Looking at such a distribution within UK Biobank shows that within the high-risk group (those with an 15-30% increased risk of developing schizophrenia assuming a 0.99 hazard ratio) the ratio of women to men is 1:1.55 (**Supplementary Note Figure 1B**). This proportion is in line with the prevalence of SCZ observed between sexes, 1:1.4. To further test whether the proportion of SCZ cases is dependent on relative height, we conducted a Pearson’s chi square test on the proportion of SCZ cases in participants with 1 standard deviation or more below the average relative height versus the proportion of SCZ cases in the rest of the participants. To make height values relative we adjusted for age and sex, resulting in an overall mean height value of 169.19 centimeter (SD = 6.17). People with more than 1 SD shorter height (< 163.02 centimeter) were grouped together and the proportion of SCZ cases in this group was compared to the proportion of SCZ cases in the remaining group (>= 163.02 centimeter). The percentage of cases was 0.31% (227 cases, 73,076 controls) in the shorter group and 0.23% (869 cases, 382,013 controls) in the taller group, and this difference was significant (χ^2^ = 17.22, *P* = 3.33 x 10^-5^).

Given these observations, a person’s expected height relative to factors that would systematically influence height, such as age and sex, is more likely to show phenotypic associations with SCZ compared to a person’s actual, or absolute, height. For this reason, we utilize relative height in our downstream analyses, that is: genome-wide association analyses of height will include relevant covariates such as sex, SES, and age.

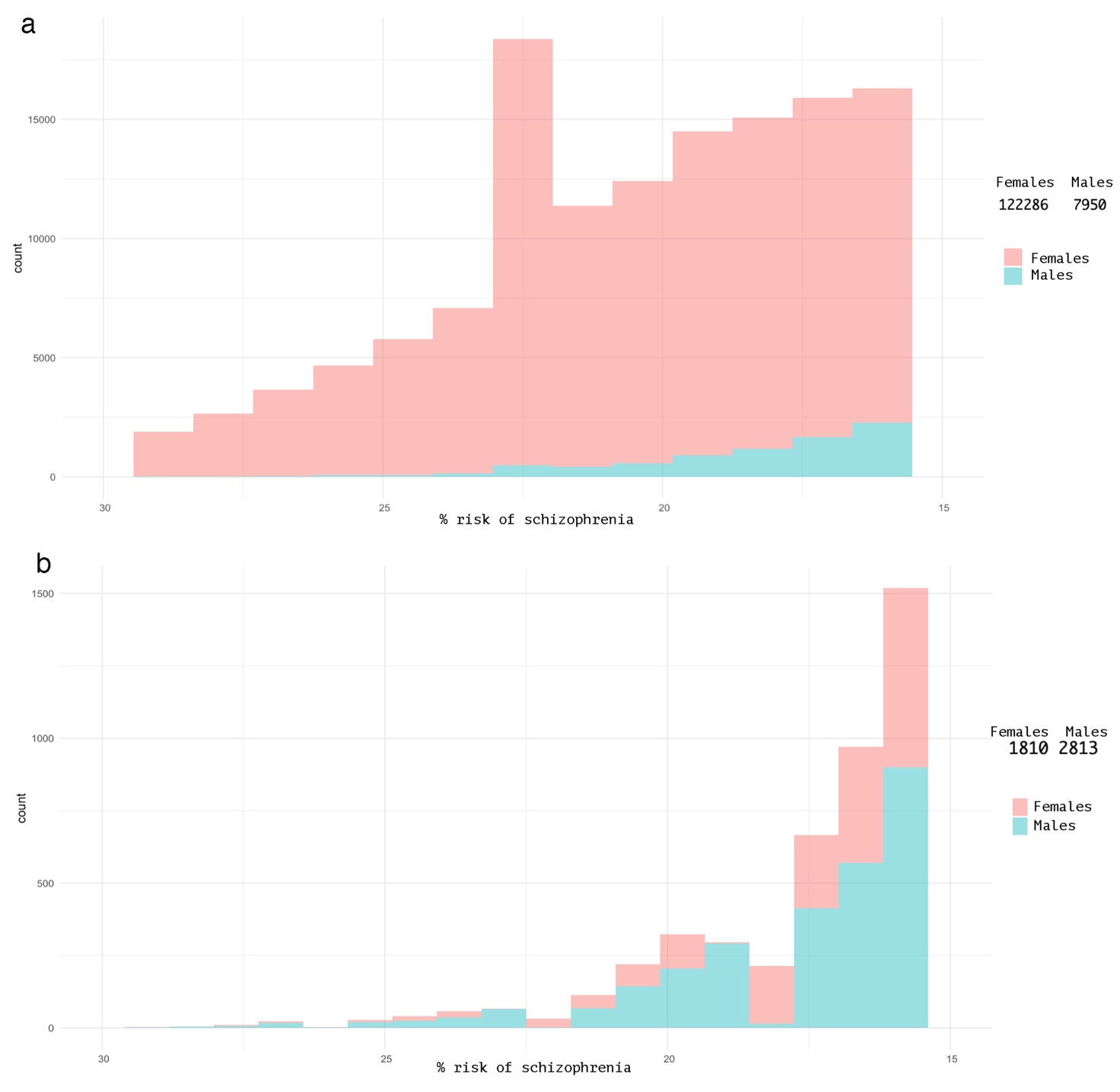

#### **Supplementary Note Figure 1. Distribution of male and female participants in the UKB at high-risk groups for SCZ under the assumption of absolute height (top) and relative height (bottom)**

*a) Female (red) to male (blue) proportion in the 145cm to 160cm group (with 15-30% increased SCZ risk assuming absolute height is related to SCZ). This group totals 122,286 females to 7,950 males and this ratio is not in accordance with the difference in prevalence for SCZ between the sexes as observed in the general populations (observed female:male ratio in the plot: 1:15; female:male ratio in the population: 1:1.4^4^). b) Female (red) to male (blue) proportion at the 15-30% increased SCZ risk group assuming that relative height is related to SCZ. This group totals 1,810 females to 2,813 males and this ratio is in accordance with the difference in prevalence for SCZ between the sexes (observed female:male ratio in the plot 1:1.5; female:male ratio in the population: 1:1.4^4^).*

### **Supplementary Note 3. Sex-stratified local genetic correlations and bivariate MiXeR**

To assess the extent of sex-specific effects within the genetic relationship between SCZ and height, we used sex-stratified GWAS summary statistics of SCZ^5^ and height (see **Supplementary** **Table 2** for an overview) to estimate and compare the significant local genetic correlations across the genome between sexes.

Using male-only SCZ and height summary statistics, 329 out of 2517 genomic regions showed significant univariate genetic signal (*P* < 1 x 10^-4^) in both SCZ and height, which is required to be able to robustly estimate a local genetic correlation. In these 329 regions, we subsequently tested the local genetic correlations between SCZ and height. Out of the 329 bivariate genetic correlations estimated, 14 regions were significant after correction for multiple testing (*α_BON_ =* α / nr. of bivariate tests conducted = 0.05/329 = 1.52 x 10^-4^; **Supplementary Note Figure 2A**). Among the 14 significant regions, 2 regions were positively correlated, while 12 regions (9 in the MHC region) showed negative correlations. The 2 positively correlated regions and 4 out of the negatively correlated regions were not seen in the combined (i.e., non-sex-stratified) analysis, and remained significant even when the same multiple testing correcting threshold was applied as in the combined analysis (*α_BON_=α/nr. of bivariate tests conducted* = .05/816 = 6.13 x 10^-5^; see **Supplementary Table 8**).

Using female-only SCZ and height summary statistics, 512 out of 2,517 genomic regions showed significant univariate genetic signal (*P* < 1 x 10^-4^) for both SCZ and height, 183 regions more than in males. Out of the 512 bivariate genetic correlations conducted, 3 survived significance correction for multiple testing (*α_BON_ = α / nr. of bivariate tests conducted* = .05/512 = 9.77 x 10^-5^; **Supplementary Note Figure 2B**). Two regions were positively correlated, and one region showed a negative correlation. The two positively correlated regions were not significant in male-analyses, while the negatively correlated region was also significant in the male analyses. All three regions were significant in the combined analysis (**Supplementary Table 8**). **Supplementary Note Figure 2C** shows the significance of female-only, male-only and combined genetic correlations within regions that were significant across sex-stratified analyses.

We next sought to see whether the 14 and 3 significantly correlated genomic regions in males and females, respectively, could be semi-replicated using the same SCZ data from the PGC but a different sex-stratified height cohort from the GIANT consortium without overlap to the UKB sample^6^. Out of the 14 genomic regions that showed significant correlation between SCZ and height in the male-only UK Biobank cohort, only 2 semi-replicated using male-only height summary statistics from the GIANT cohort (*α_BON_ =* α / nr. of significant correlated regions = 0.05/14 = 3.57 x 10^-3^; **Supplementary Figure 2D**). For females, none of the 3 significantly genetically correlated regions observed in UK Biobank female-only data, semi-replicated using female-only height summary statistics from the GIANT consortium (*α_BON_ =* α / nr. of significant correlated regions = 0.05/3 = 1.67 x 10^-2^). As one of the semi-replicated regions in males was not significant in the combined analysis, this adds robust evidence for only a small male-specific effect in the SCZ-height relation. However, it should be noted that the sample sizes for the sex-stratified SCZ GWAS summary statistics is larger for males (N = 68,287, N-cases = 33,097) than for females (N = 54,513, N-cases = 17,710), hence the power to detect sex-specific effects is not equal between the sexes. Future investigation of sex-specific genetic effects would benefit from larger and equally sized sex-stratified GWAS summary statistics. A complete overview of the results from all analyses described in this **Supplementary Note 4** can be found in **Supplementary Table 8**.

Sex-stratified bivariate MiXeR was used to see whether the proportion of estimated shared causal SNPs was different for males and females. MiXeR was conducted in sex-stratified GWAS datasets and showed ~800 (SE = 200) shared variants for males, and ~700 (SE = 300) shared variants for females. Sign-concordance for male shared variants was 49% (SE = 1%), and 57% (SE = 7.6%) for female shared variants. AIC fit indices were positive for both models, while BIC was only significant for the male model, indicating slightly better fit in the male models likely due to larger SCZ GWAS sample size. These results align with sex-stratified local genetic correlation analysis in that there is little evidence for a sex-specific genetic relationship between SCZ and height.

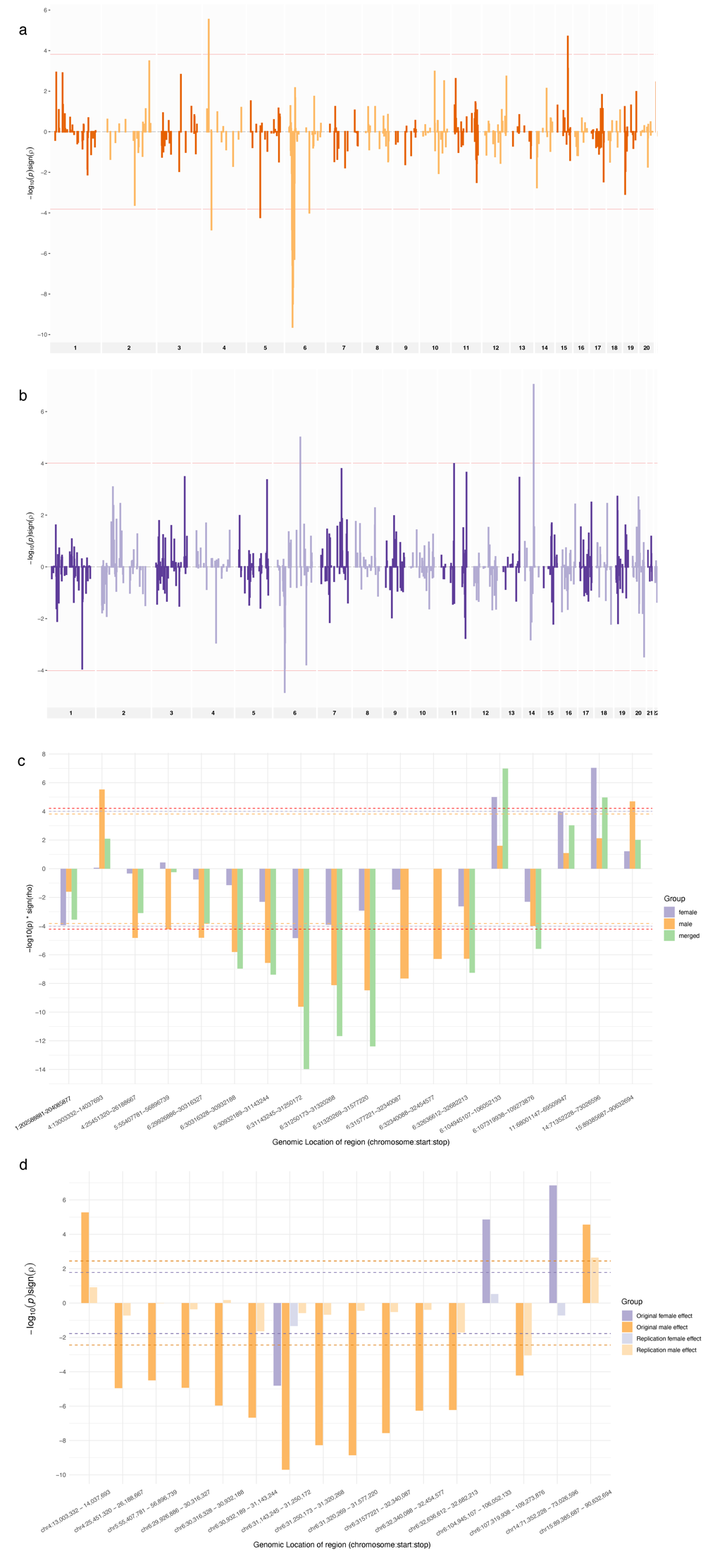

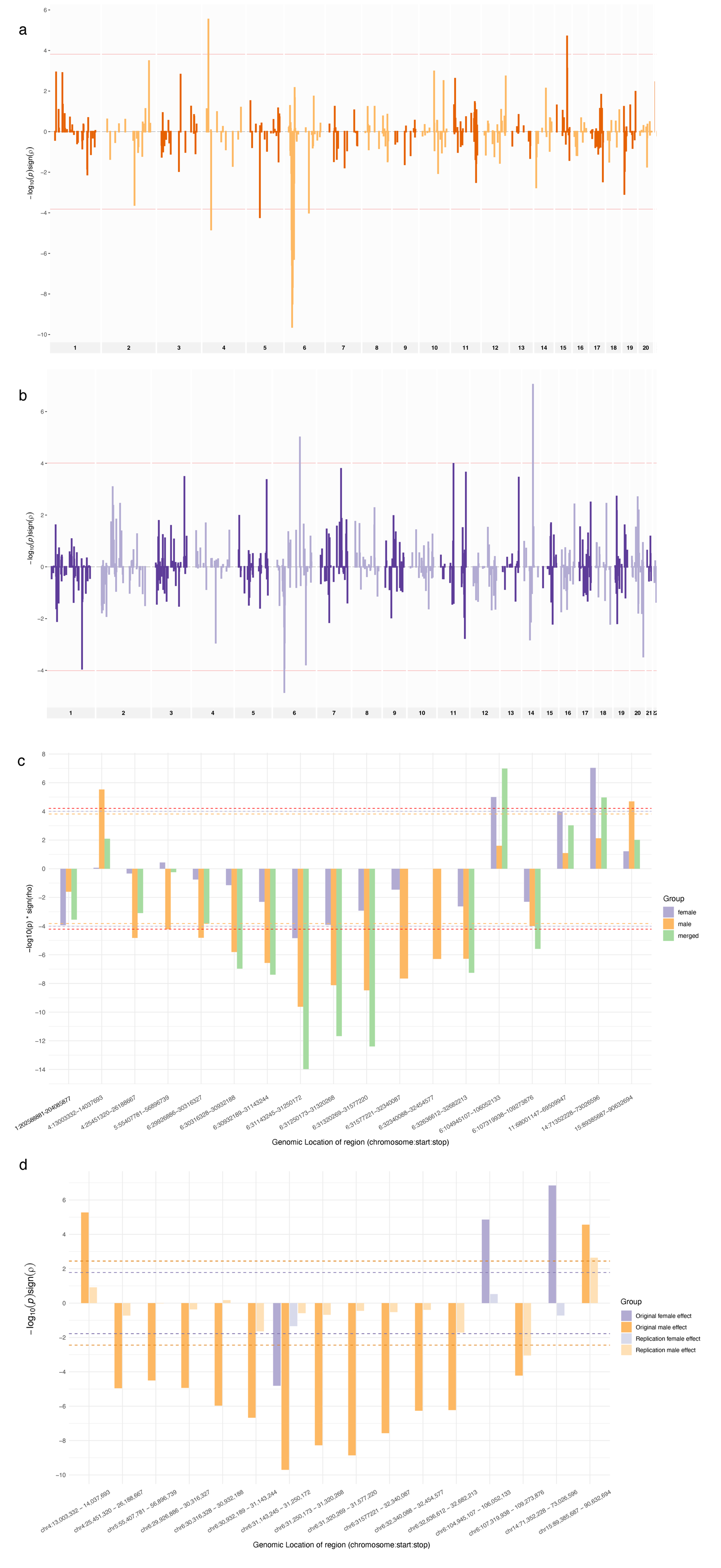

**Supplementary Note Figure 2. Sex-stratified local genetic correlations between SCZ and height**

*a) All (386) estimated genetic correlations between SCZ and height in the male sample, with genomic location on the x-axis and the sign-weight (+ / -) -log10 of the p-values corresponding to the correlations on the y-axis (red dotted line indicating the genome-wide significance threshold correcting for the 386 conducted tests for positive (top) or negative (bottom) correlation estimates, respectively). b) Same figure for the female sample, with a total of 512 test conducted and corrected for. c) All genomic regions from the analyses in a and b that were significant in males (orange bars) and/or females (purple bars) along with the effect from the combined analysis (green bars). Orange, purple, and red lines indicate significance thresholds correcting for number of tests from male-regions (386), female-regions (512), and combined regions (816), respectively. d) Assessment of sex-stratified genetic effects in regions significant in the UKB sample (dark orange/purple) compared to the effect seen in sex-stratified data from the GIANT consortium (light orange/purple). Only two regions in males replicated in the GIANT sample.*

### **Supplementary Note 4. Linearity of association of SCZ with height using stratified height data**

The genetic association between SCZ and height could potentially be non-linear, i.e., the strength of the relationship could differ between various strata of height values. Specifically, it is possible that the genetic determinants of extreme lower or higher ends of the height distribution are primarily associated with SCZ as opposed to a general, linear tendency throughout the height continuum. Estimating whether certain strata, or bins, of height are more associated with SCZ is difficult due to power limitations induced 1) by splitting an overall sample into smaller sub-samples, and 2) by decreasing the range of phenotypic values on which to detect SNP-associations (i.e., restriction of range).

To overcome this limitation of assessing the linearity of the genetic relationship between SCZ and height, we used a cumulative trend approach where separate GWASs were conducted of the 10%, 20%, 30%... 90% subjects in the *lower* end of the height distribution in UKB, separately, and local genetic correlations were estimated with the full SCZ summary statistics. We then repeated the analyses, but instead conducted GWASs on the 10%, 20%, 30%... 90% subjects in the *higher* end of the height distribution and estimated the genetic correlations with SCZ. Trends in local genetic association were then compared between both approaches to establish potentially different contributions to the local genetic SCZ-height correlations at specific strata of the height spectrum. A control trend was added where a random sample of 10%, 20%, 30%...90% height values were gathered for separate GWASs to allow a ‘baseline’ comparison. The regions tested where the same as the 9 showing robust local genetic correlation in the analysis of the full-sample of European ancestry.

Based on a descriptive analysis of the trend results, there was but one clear deviation in the cumulative genetic association within the genetic regions correlated between SCZ and height that could be distinguished for genetic trends starting at the tallest vs. shortest subject (**Supplementary Note Figure 4**). In region 107,319,938 - 109,273,876 on chromosome 6, genetic associations based on the lower end of the height distribution showed a much stronger rise in significance, surpassing that of the randomly drawn reference group after 50% of the subjects within the lower end of the height distribution was analyzed. The genetic association in this region that was based on subjects in the higher end of the height distribution showed essentially no sign of association until more than 80% of the subjects were included. This suggests that for this region, the local genetic correlation only becomes significant when the 20% shortest individuals are included in the analysis (see blue line in 9^th^ panel of **Supplementary Note Figure 4**), and thus, the genetic correlation between SCZ and height is predominantly between SCZ and the 20% shortest individuals in this locus. In the other 8 genomic regions assessed, there was little sign of deviation between the association within the lower and higher end of the height distribution and at no point did their significance clearly surpass the randomly drawn reference condition, suggesting that the association between SCZ and height is linear across the height strata within these regions. Summarizing, extreme short or tall stature thus not seem more genetically associated with SCZ than an overall drawn distribution of height measurement values (see **Supplementary Table 10** for all results).

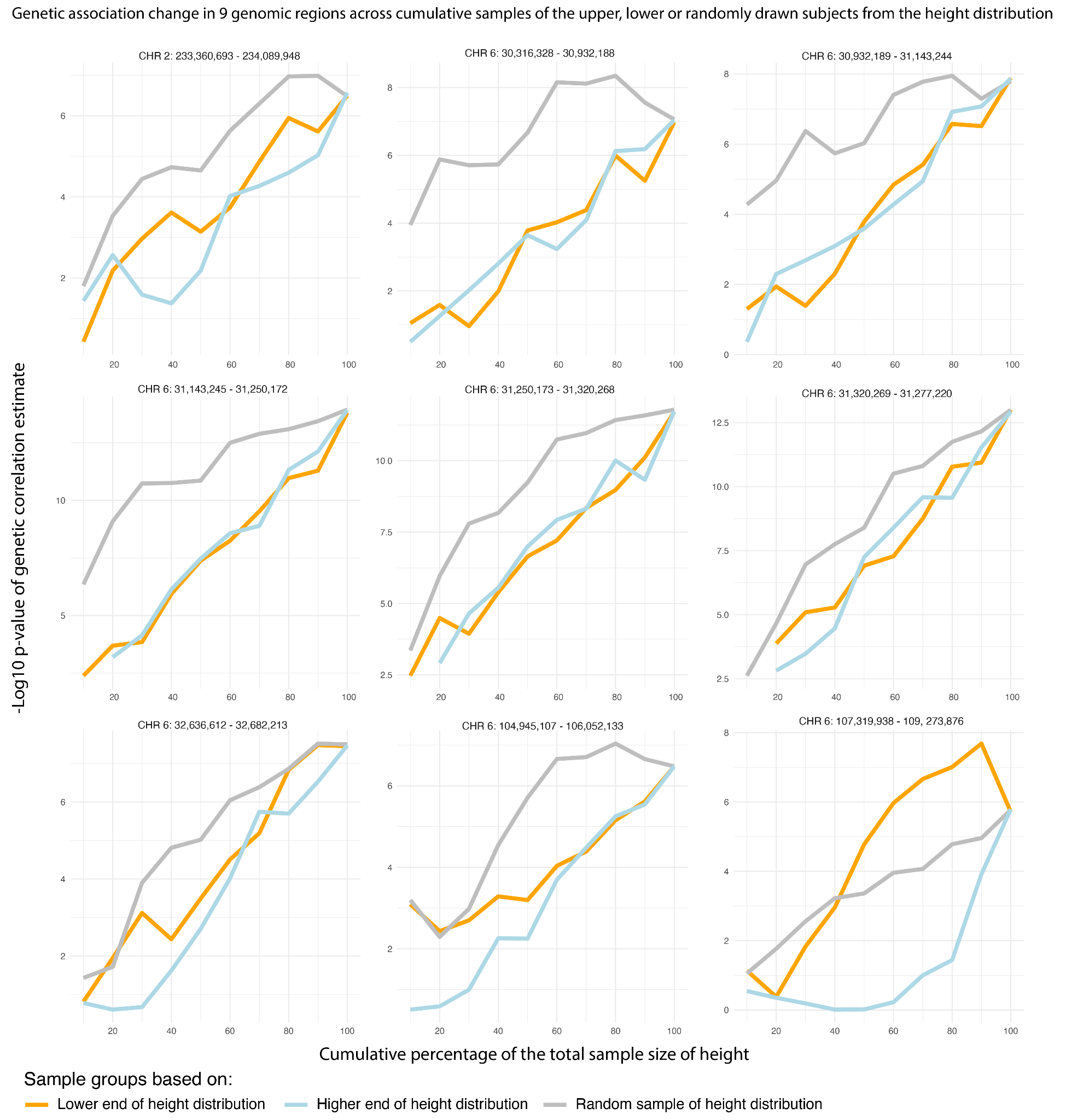

#### **Supplementary Note Figure 3. Assessment of linearity in associations between SCZ and strata of height**

*Comparison of associations between 10%, 20%, … 90% subjects from the lower end of the height distribution (orange), higher end of the height distribution (light blue), and random selection of height values (grey) across 9 genomic regions. -Log10 p-value of the local genetic correlation between SCZ and height at each cumulative stratum of height is plotted on the y-axis and cumulative percentage of subjects included in the different height strata GWASs are plotted on the x-axis. Grey trend lines (random sample of height distribution) are systemically higher (more significant) than blue (higher end distribution trend) or orange (lower end distribution trend) due to estimating genetic association between SCZ and a larger phenotypic height range (i.e., no restriction of range in the height data of the random sample). Upper and lower end of height distribution trends are close to indistinguishable in 8 genomic regions, except for chromosome 6 107,319,938 – 109,273,876 (region 1052; lower right panel) where the lower end of the distribution seems to be driving the genetic association with SCZ, i.e., the blue line only nears significance when the last 20% shortest participants are included in the local genetic correlation estimate.*

### **Supplementary Note 5. Assessing genetic correlations between SCZ and height using a within-sibling GWAS on height**

Genetic correlations have been shown to be sensitive to indirect genetic effects, such as confounding by assortative mating, cryptic relatedness through population stratification, and gene-environment interactions^7^. Different family-designs, such as within-sibling genome-wide association studies, are less sensitive to such confounding resulting from indirect genetic effects and can more reliably estimate direct SNP association effects with phenotypes of interest. Using a within-sibling GWAS on height^7^ (N_sibling-pairs_ = 75,030), we sought to estimate the genetic correlation between SCZ and height and see whether significant genomic regions replicated. Although statistical power was lower in the within-sibling GWAS due to the smaller sample size (N=75,030) compared to the overall UK Biobank GWAS (N = 382,754), 4 out of 9 genomic regions showed a significant genomic correlation between SCZ and height using the within-sibling height GWAS (*α_BON_ = α / nr. of significant correlated regions* = 0.05/9 = 5.56 x 10^-3^; **Supplementary Note Figure 5**; **Supplementary Table 11**). An additional 5^th^ region of the 9 was also significant; however, the sign of the correlation was flipped from being positive to negative, possibly suggesting strong influences of both direct and indirect genetic effects in this region. These results suggest that, at least in these 4 regions, the genetic relationships between SCZ and height are based on direct genetic effects, and not driven by cryptic relatedness, assortative mating, or gene-environment interactions. The 5 remaining genomic regions might lack statistical association due to lowered statistical power or be (partly) driven by indirect genetic effects.

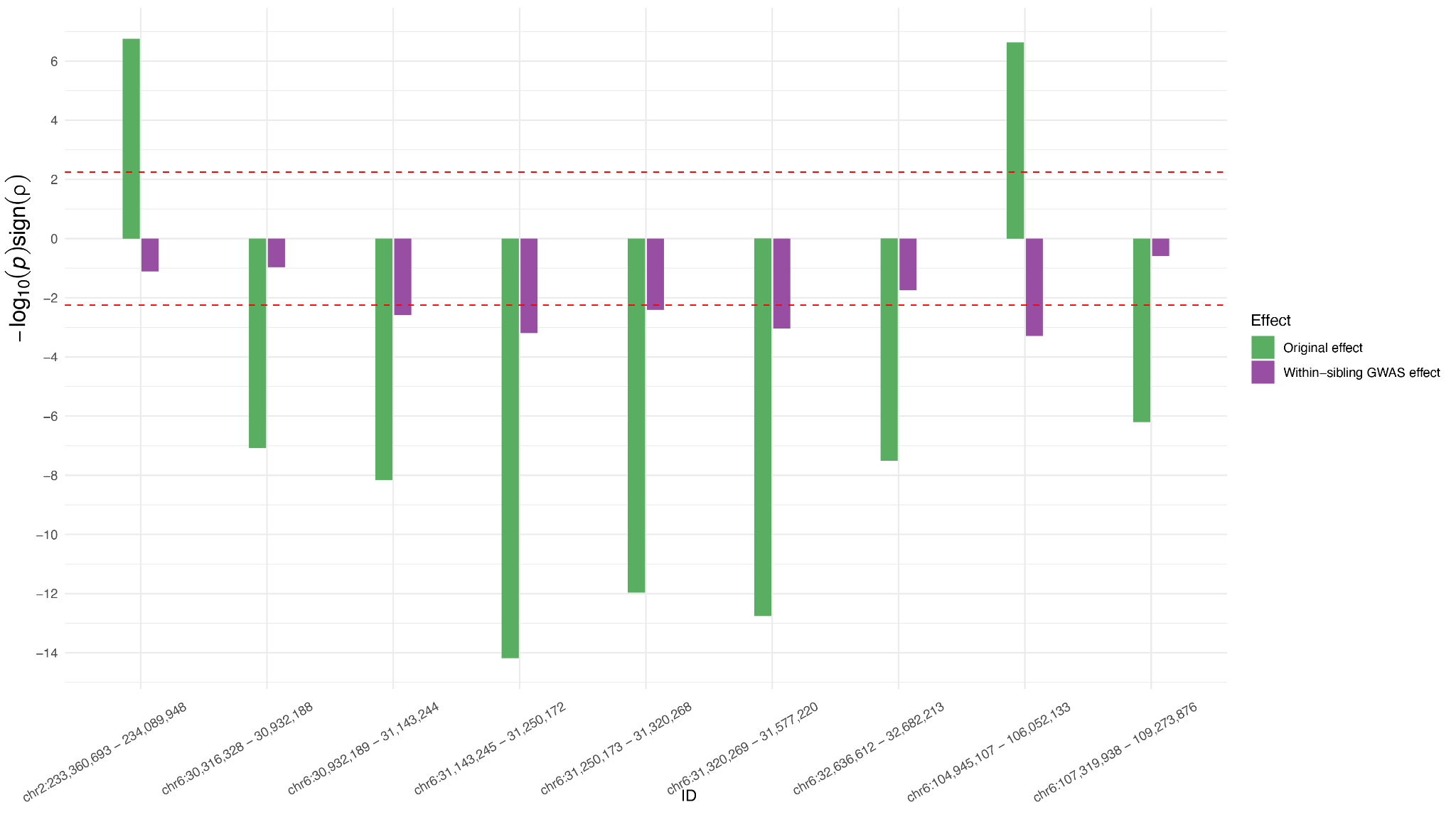

#### **Supplementary Note Figure 4. Replication of the 9 local genetic correlations using a within-sibling GWAS on height to assess the influence of indirect genetic effects**

*Local genetic correlations were computed between SCZ and height using a within-sibling GWAS on height within the 9 genomic regions showing significant local genetic correlations between SCZ and height in European ancestry. Original European effects are shown in green bars and effects based on the within-sibling GWAS of height are displayed in purple. Genomic regions are plotted on the x-axis and the y-axis represents the sign-weight (+ / -) -log10 P-value of the correlations. The SCZ-height local genetic correlations were replicated in 4 out of 9 regions using the within-sibling analyses suggesting that the genetic effects within these regions are based on direct (as opposed to indirect) genetic effects.*

### **Supplementary Note 6. Fine-mapping of genomic regions correlations between SCZ-height**

Regional or local genetic correlations between traits are likely driven by genetic effects impacting the same genes, which downstream have an impact on the trait’s measurement value. Between SCZ and height, we observed 9 significantly correlated genomic regions. We used FLAMES^8^ to conduct fine-mapping of the 9 genomic regions to identify the most likely effector genes (i.e., those genes that most likely mediate a genomic locus association with a trait) underlying the local SCZ-height genetic correlation (**Methods**). SCZ and height converged on the same prioritized effector gene in three loci. In region 410 (chromosome 2 233,360,693 - 234,089,948), *GIGYF2* was the top prioritized gene for SCZ (FLAMES score = 0.11) and height (FLAMES score = 0.045). In region 974 (chromosome 6 31,250,173 - 31,320,268, subsection of the MHC region which included multiple genetic correlations), *HLA-C* was the top prioritized gene for both SCZ (FLAMES score = 0.06) and height (FLAMES score = 0.10). In region 1050 (chromosome 6 104,945,107 - 106,052,133, outside the MHC region), *LIN28B* was the top prioritized gene for both SCZ (FLAMES score = 0.14) and height (FLAMES score = 0.34). For a complete overview of the top prioritized genes across all 9 genomic regions, see **Supplementary Table 12**.

GIGFY2 (Grb10-Interacting GYF Protein 2) is related to growth and developmental processes through its implication with the insulin/IGF-1 signaling pathway, which is specifically crucial for neurodevelopment and synaptic plasticity^9^. *HLA-C* (Human Leukocyte Antigen C) is central to immune system regulation and response and expressed broadly in cell types like nucleated cells, immune cells, epithelial cells, and placental cells^9^. *LIN28B* (Lin-28 Homolog B) is a regulator of microRNA, of growth patterns during early development, and of neuronal differentiation and maturation, and is associated with metabolism and puberty age-of-onset. *LIN28B* is highly expressed in the pituitary^9^, the only tissue type that was significantly annotated to both SCZ and height.

### **Supplementary Note 7. Derivation for conditional local genetic analysis in LAVA**

The aim of conditional local genetic analyses is to assess whether the genetic covariance between two traits within a genomic locus changes when conditioned on the shared genetic covariance of an external trait (or *covariate*). If the conditional covariance is different from the marginal covariance, this implies that the genetic signal of the covariate trait overlaps with the genetic signal of the main traits, and this might further our understanding of the nature of the genetic covariance observed between the two main traits. The derivation of a statistical test that can assess the difference between a marginal and conditional effect is given as follows:

$$\text{For predictors }X_{1}, X_{2} \text{and outcome} Y:$$

$\text{Writing }C\text{ for covariances and }V\text{ for variances, and with determinan}$t $D=V_{1}V_{2}-C_{12}^{2}$ $\text{for the predictor covariance matrix:}$

$$\text{Marginal effects:} \beta_{1}=\frac{C_{1Y}}{V_{1}} \text{and} \beta_{2}=\frac{C_{2Y}}{V_{2}}$$

$$\text{Conditional effects: }\beta_{1|2}=\frac{1}{D}\left( V_{2}C_{1Y}-C_{12}C_{2Y} \right) \text{and} \beta_{2|1}=\frac{1}{D}\left( V_{1}C_{2Y}-C_{12}C_{1Y} \right)$$

For $X_{1}$, testing $H_{0}:\beta_{1}=\beta_{1|2}$, or equivalently $H_{0}:\beta_{1}-\beta_{1|2=0}$. If the marginal and conditional effects exist, this implies that $V_{1}\neq0$ and $D \neq0$ (the latter will be false if ${cor\left( X_{1},X_{2} \right)}^{2}=1$, i.e., complete collinearity), and as such we can multiply by these.

We therefore have:

$$(\beta_{1} - \beta_{1|2})V_{1}D=\left( V_{2}C_{1Y}-C_{12}C_{2Y} \right)V_{1}-\left( V_{1}V_{2}-C_{12}^{2} \right)C_{1Y}$$

$$=V_{1}V_{2}C_{1Y}-V_{1}C_{12}C_{2Y}-V_{1}V_{2}C_{1Y}+C_{12}^{2}C_{1Y})$$

$$=C_{12}^{2}C_{1Y}-V_{1}C_{12}C_{2Y}$$

$$=C_{12}\left( C_{12}C_{1Y}-V_{1}C_{2Y} \right)$$

And as such, $\beta_{1}-\beta_{1|2}=-\frac{1}{V_{1}}C_{12}\frac{\left( C_{12}C_{1Y}-V_{1}C_{2Y} \right)}{D}=-\frac{1}{V_{1}}C_{12}\beta_{2|1}$, i.e., the difference will be zero either if $C_{12}=0 \text{or} \beta_{2|1}=0$. In other words, the conditional effect of $X_{1}$ will equal its marginal effect if either $X_{1}$ and $X_{2}$ are independent, or the conditional effect of $X_{2}$ on $Y$ given $X_{1}$is zero. If $X_{1}$ and $X_{2}$ are dependent ($C_{12}\neq0$), then the significance of $\beta_{2|1}$ is equal to the significance of the difference between $\beta_{1}and \beta_{1|2}$.

### **Supplementary Note 8. Mendelian randomization analysis between SCZ and height**

To assess the direction of phenotypic association between SCZ and UKB-height, we used Mendelian randomization. GWAS summary statistics were clumped to a set of independent lead SNPs, using the 10K EUR reference panel as LD reference. A total of 4,352 clumped SNPs were further used to construct an LD correlation matrix used for the Mendelian randomization analysis. Out of the 4,352 clumped SNPs, 171 showed significant association with SCZ and not UKB-height (at *P* < 5 x 10^-8^), and are therefore potential instrumental variables. A HEIDI filter was subsequently applied to correct for still existing pleiotropy, resulting in 169 independent instruments that were used to assess the phenotypic effect of SCZ on height. For UKB-height, 2,249 out of the 4,352 clumped SNPs were significantly associated with UKB-height and not SCZ (at *P* < 5 x 10^-8^), and after HEIDI filtering, 2,155 independent instruments remained. These 2,155 independent instruments were used to assess the phenotypic effect of UKB-height on SCZ. The results yielded more evidence for height having a causal effect on SCZ (Height causing SCZ: standardized *β_height_* = -0.024, SE = 0.006, *P* = 1.99 x 10^-5^; SCZ causing height: standardized *β_SCZ_* = -0.02, SE = 0.008, *P* = 0.02). For our conditional analyses in LAVA, we therefore choose to use SCZ as the outcome variable and height as the predictor.

### **Supplementary Note 9. Replication of SCZ-height shared SNPs, genes, and functional annotations in non-EUR ancestry**

To assess the extent to which the shared SNPs, genes and biological annotations between SCZ and height in EUR-ancestry GWAS is also present in non-EUR ancestry, we sought to replicate the lead SNP, gene, and gene-annotation findings in SCZ^5^ and height^10^ GWAS based on East-Asian (EAS), African (AFR) and Latino (LAT) ancestry (**Methods)**. Global SCZ-height genetic correlations were near-zero and non-significant for EAS (*r*_g_ = -.00, SE = .03, *P* = .99) and LAT (*r*_g_ = -.10, SE = .01, *P* = .23); however for AFR, the genetic correlation between SCZ and height was positive (*r*_g_ = .29, SE = .13, *P* = .02), indicating potentially ancestry-dependent genetic relationships between SCZ and height. Only analyses in East-Asian ancestry showed significant associations among EUR lead SNPs (2/22 significant in both SCZ and height; **Supplementary Table 3**), gene-based analysis (9/142 significant in both SCZ and height; **Supplementary Table 4**) and gene-property analysis (pituitary tissue significant in both SCZ and height; thyrotropic cells significant for SCZ and mesenchymal stem cells significant for height; **Supplementary Table 7**). Lack of overlap between SCZ and height based on African or Latino ancestry could signify a potential ancestry-specific relationship. Indeed, phenotypic data from the UKB showed that the age and sex-adjusted mean height was on average slightly *higher* for the SCZ-cases compared to controls with AFR and LAT ancestry (**Supplementary Note Table 2**); however, these are only descriptive results that cannot be tested formally as the sample sizes are far too small. Similarly, inability to replicate genetic effects in SCZ and height based on AFR and LAT cohorts could be due to the lower sample size and thus lack of statistical power in these GWASs compared to EAS cohorts.

#### **Supplementary Note Table 2:** phenotypic sex/age adjusted height measurements from the UKB between SCZ cases and controls across different ancestries

|  | Height difference | Mean Height in cm | SD Height in cm | N |
| --- | --- | --- | --- | --- |
| AFR SCZ | .57cm | 168.02 | 6.22 | 91 |
| AFR Control |  | 167.59 | 6.21 | 8,168 |
| EAS SCZ | -2.21cm | 160.18 | 2.37 | 8 |
| EAS Control |  | 162.39 | 6.93 | 2,404 |
| LAT SCZ | .49cm | 167.43 | 6.93 | 23 |
| LAT Control |  | 166.94 | 6.39 | 3,628 |

### **Supplementary Note 10. Attempts to replicate genetic correlations in non-European ancestry**

After establishing significant local genetic correlations between SCZ and height in data from subjects with European ancestry, we sought to replicate these significant genomic regions in GWAS summary statistics available for non-European ancestry. GWAS summary statistics with sufficient power (N > 20,000) were available for East-Asian ancestry through the GIANT consortium^11^ for height and the PGC for SCZ^5^.

Eleven genomic regions (based on East-Asian LD structure) showing overlap with the 9 genomic regions harboring significant correlations in the European data, were tested for bivariate genetic correlations in the East-Asian data (**Methods**). None of the local genetic correlations in these 9 genomic regions surpassed the significance threshold after correction for multiple testing (*α_BON_ = α / nr. of significant correlated regions in European data* = 0.05/9 = 5.56 x 10^-3^; **Supplementary Note Figure 5**; **Supplementary Table 14**). That is, we were unable to replicate significant genetic correlations observed in European samples in East-Asian ancestry. While this could be due to obvious differences in sample sizes and thus statistical power between the European and East-Asian cohorts, it might also suggest ancestry-specific differences in genetic effects (and possibly in the phenotypic SCZ-height relation, which has to the authors’ knowledge yet only been established in samples of European ancestry). As such, interpretation of the causes for the genetic relationship between SCZ and height might be specific to European genetic ancestry.

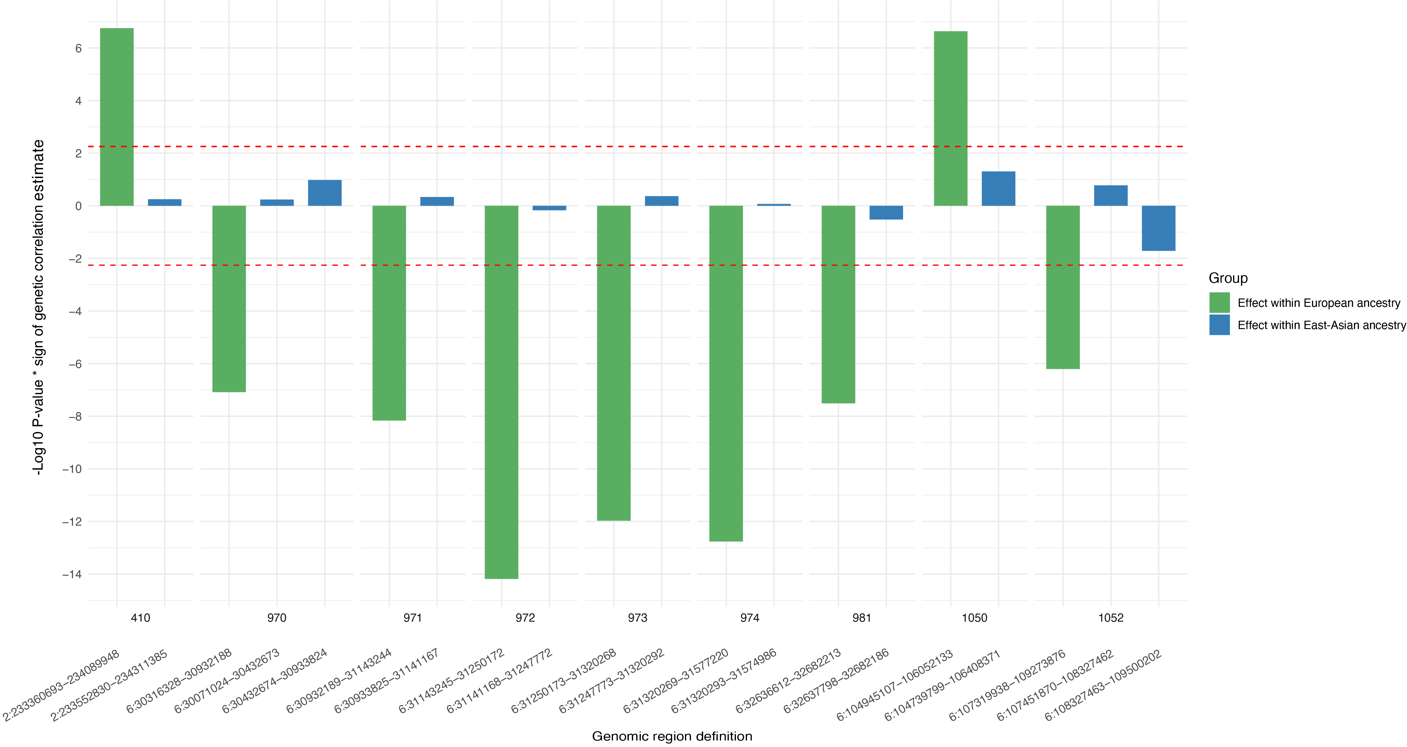
**Supplementary Note Figure 5. Local genetic correlations seen in European ancestry compared to East-Asian ancestry**

*All 9 local genetic correlations originally observed in EU ancestry (green) and then tested in EAS ancestry (blue) with genomic location on the x-axis and the sign-weight (+ / -) -log10 of the p-values corresponding to the correlations on the y-axis (red dotted line indicating the replication significance threshold correcting for the 9 conducted tests). Local genetic correlations were computed using SCZ and height GWAS summary statistics from East-Asian ancestry within genomic regions that overlap with regions significantly correlated in European ancestry. In none of the 9 genomic regions do the East-Asian local genetic correlations between SCZ and height approach significance.*

### **Supplementary Note 11. Summary and discussion of all overall and region-specific findings**

#### *Overlapping genome-wide results between SCZ-height*

To summarize across all analyses conducted in general, we find 142 shared genes between SCZ and height. These genes show significant enrichment for genomic positional gene-sets as well as gamma-interferon signaling, a gene-set involving immune response. Gene property analysis find the pituitary as a tissue that is enriched for both expression of SCZ and height genes. Further inspection finds that within the pituitary, the mesenchymal cells to be enriched for height genes and thyrotropes to be enriched for SCZ genes. This indicates different functions of the pituitary as relevant for SCZ and height: mesenchymal cells are more involved in maintenance and structural integrity of the pituitary, while thyrotrope cells are primarily related to the regulation of thyroid hormone in the body, a hormone found to be negatively associated with early psychosis, but positively associated with patients with chronic SCZ^12^. If the genetic effect on thyrotrope cells and thyroid levels is a main component of the SCZ-height relation, that could explain the mix of positive and negative local genetic correlations, as both increases and decreases in thyroid hormone levels are associated with SCZ diagnosis, while height is only positively associated with thyroid hormones^13^.

Though SCZ and height are enriched in different cell-types in the pituitary, it is highly likely that the proper functioning of each cell-type relies on the functioning of the other, i.e., if mesenchymal cell functions are dysregulated, this will likely lead to problems in maintaining the extracellular environment in the pituitary which indirectly will influence thyrotrope cells. Conversely, mesenchymal cells have been found to express a thyroid hormone receptor and have functions like maintenance and differentiation depend on thyroid signaling.^14^

In terms of genomic region-specific effects, we will go through all results region by regions among those that show robust genetic correlation between SCZ and height.

#### *Region 410 chromosome 2 233360693 — 234089948*

This region shows a positive genetic correlation between height and SCZ. None of the covariates tested in conditional LAVA analysis? showed significant effects within this region. Here, *GIGYF2* was fine-mapped and prioritized to both SCZ and height. *GIGYF2* is a gene related to a microRNA complex that plays important roles in silencing mRNA during mammalian development. Expression of *GIGYF2* is highest from week 8-17 post conception, with broad expression across tissues^9^. *GIGFY2* has also been shown to repress cytokine translation when bound to the complex with *EIF4E2*, which can lead to prevention of anti-inflammatory cytokines being released and as a result potentially increased inflammation in response to viral pathogens^15^.

One explanation for why no significant covariates were found for this region could be that the genetic covariance between SCZ and height is primarily involved in early development, such that using GWAS summary statistics of covariates measured in adult participants will not reflect relevant genetic effects within this locus.

#### *Regions 970-974, 981 chromosome 6 30316328 —31577220 & 32636612—32682213*

As these regions are all in the MHC and show concordant results across multiple analyses, we will discuss the findings pertaining to them collectively. These regions are within the MHC, an important region of immune system regulation and functioning. This region is challenging to study due to being very gene-dense and showing complex long-range LD structure. Nevertheless, we find that the genetic signal reflected in baseline white blood cell count significantly overlaps with the genetic covariance shared between SCZ and height in these regions. Moreover, despite the high density of genes, fine-mapping prioritized the same gene, *HLA-C* out of 78 genes, as the most likely effector gene for both SCZ and height. The *HLA-C* protein is expressed as an antigen binding motif for viral peptides by almost all cells in the human body and is used by immune cells (lymphocytes and monocytes) to recognize and destroy infected cells. Individual HLA genetic variation may explain differences in immune responsivity to virus across individuals, and activity of *HLA-C* can increased white blood cell production and lead to increased levels of inflammation^16^. We also find phenotypic evidence for a higher white blood cell count for people with SCZ and short stature alike, lending evidence to a potential role of immune regulation and inflammation in connecting SCZ and height genetically. Speculating, high baseline levels of white blood cell count might be protective against infectious disease at the cost of more inflammation in the body, that during development might be a risk factor for both SCZ and shorter stature.^17–19^

#### *Region 1050 chromosome 6 104945107 —106052133*

This region shows a positive correlation between SCZ and height, while the estimate changes to negative when correlations are estimated between SCZ and a within-sibling GWAS of height, indicating potentially both indirect and direct genetic effects influencing SCZ and height in this locus. Similar to region 410, none of the covariates tested showed significant effects within this region, but fine-mapping confidently associates *LIN28B* to both SCZ and height. *LIN28B* is highly expressed in pituitary cells and has large effects on both mesenchymal and thyrotrope cell development. Overexpression of *LIN28B* has been shown to be present in thyrotrope precursor cells, indicating the involvement of *LIN28B* in controlling thyrotrope cell pools^20^. In mesenchymal cells, *LIN28B* induces differentiation of mesenchymal cells and promotes mesenchymal protective effects for the microenvironment^21^. *LIN28B* KO zebra fish models also show stunted growth and brief overexpression of *LIN28B* have lasting increasing effects of growth^22,23^. Lastly, *LIN28B* has also been shown to play a role in the fetal development of lymphocytes and overexpression of *LIN28B* is associated with inflammation markers in the body.^24,25^

#### *Region 1052 chromosome 6 107319938 —109273876*

This region shows a negative, non-linear association between SCZ and height and is mediated by the shared genetic covariance with intra-cranial volume, suggesting that genetics involved in shorter stature and SCZ are related through involvement of the development of the cranium, and potentially also brain development. Fine-mapping results did not convergence on effector genes, however, height prioritized gene *FOXO3* has previously been inversely associated with SCZ and intracranial volume^26^, suggesting that the actual effector gene in this region might be *FOXO3* for SCZ.

These findings point to a complex genetic relation between SCZ and height, that converges on biological processes involving immune response sensitivity and the pituitary, a gland tightly associated with immune functions^27^. Gene enrichment analyses on cell types within the pituitary found thyrotropic cells (TC) and mesenchymal cells (MSC) to be associated with SCZ and height, respectively. These cells interact by mesenchymal differentiation and function being activated by thyroid stimulating hormone secreted from TCs^28,29^. MSCs are further involved in anti-inflammatory processes by responding to inflammation factors and migrating to and promoting tissue repair in inflamed areas^30,31^. Thyroid hormones increase proliferation and activity of immune cells, such as T-cell lymphocytes, and inflammation causes in turn breakdown of thyroid hormone^32–34^. Indeed, the genetic signal of lymphocyte cell count was shown to underly the genetic correlation between SCZ and height in multiple genomic loci within the MHC region. Lymphocytes respond to *HLA-C* presentation by infected cells and utilize interferon gamma signaling (one of the significantly associated gene-sets) to enhance cytotoxic activity and cell-destruction, increasing inflammation in the affected area^35^ (see **Supplementary Note Figure 6** for a detailed schematic overview).

Among the prioritized genes, *LIN28B* is mainly expressed in the pituitary and testis, exhibits effects on growth and developmental timing^22,23^, and regulates the development of TCs^20^, MSCs^21^ and immune cells^24^. *GIGYF2* and *HLA-C* are involved in pathways related to the regulation of immune response to pathogens^15,16,36^. Polymorphisms in *GIGYF2*, *LIN28B*, and *HLA-C* could potentially lead to variation in baseline inflammation. Presence of more inflammation and dysregulated thyroid hormone levels in the body, features that are often seen in patients with SCZ^12,37^, can affect linear growth processes during development^38–40^ and lead to average differences in height between SCZ-cases and controls. If the genetic effect on TCs and thyroid levels is a main component of the SCZ-height relation, that could explain the mix of positive and negative local genetic correlations, as decreased thyroid-stimulating hormone is associated with first-episode psychosis and increase in thyroid-stimulating hormone is seen after multiple psychotic episodes^12^. This is in contrast to height which only positively associates with elevated thyroid-stimulating hormone^13^.

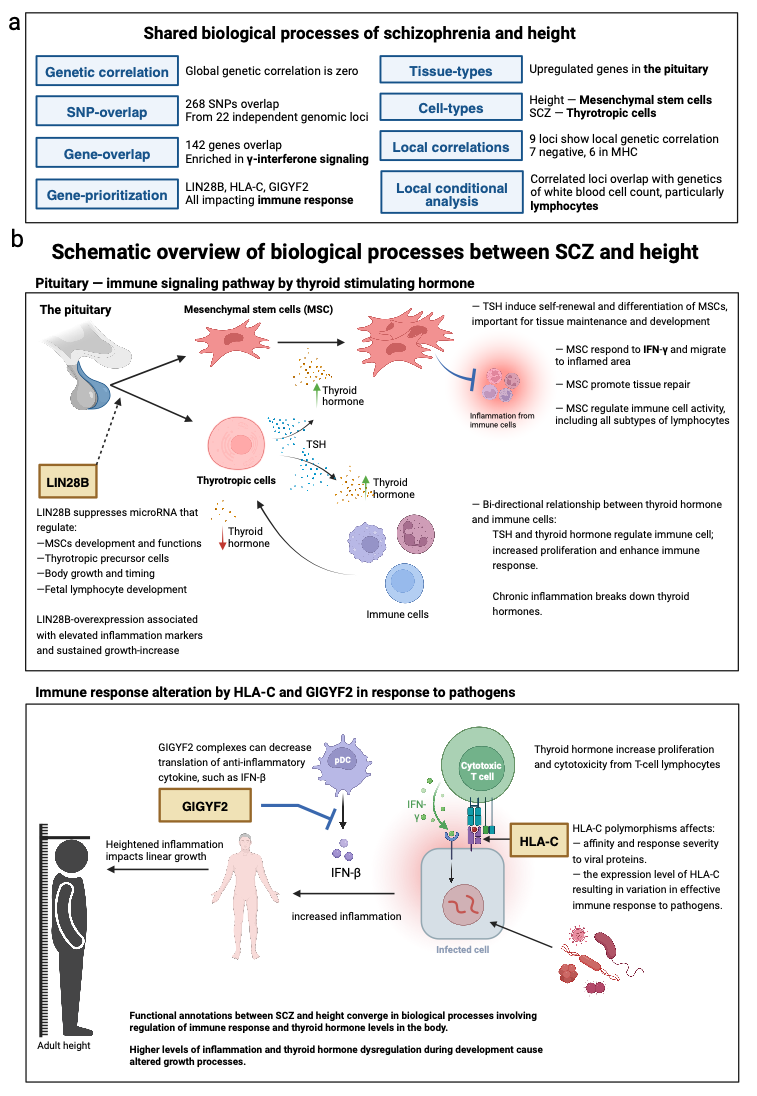

**Supplementary Note Figure 6. Overview and putative hypothesis on how SCZ and height are related**

*a) Summary of results from genome-wide and local genetic correlation analyses; b) Schematic overview of functional convergence in ‘pituitary—immune signaling pathway by thyroid stimulating hormone’ and ‘immune response alteration by HLA-C and GIGYF2 in response to pathogens’ based on annotations significant for both SCZ and height. Genes in yellow boxes (GIGYF2, LIN28B, and HLA-C) represent overlapping genes between SCZ-height from gene-prioritizing with FLAMES. Summarizing: within the pituitary, thyrotrope cells are enriched for SCZ and mesenchymal cells for height. Thyrotrope cells influence mesenchymal cell differentiation and function through secretion of thyroid hormone*^28,29^*. Mesenchymal cells respond to cytokines secreted by immune cells and are involved in anti-inflammatory processes*^30,31^*. Thyroid hormones also promotes differentiation and function of immune cells, and resulting inflammation promotes the turnover of thyroid hormone*^32–34^*. LIN28B acts in the pituitary by regulating cell-precursors to thyrotrope cells, and the development of mesenchymal and immune cells*^20,21,24^*. GIGYF2 and HLA-C are involved in pathways related to regulation of immune response to pathogens, and in sum, genetic polymorphisms in LIN28B, GIGYF2 and HLA-C could result in variation in individuals’ immune responsivity*^15,16,36^*. When thyroid signaling and immune response are dysregulated, as is often seen in patients with SCZ*^12,37^*, this can lead to impairment of growth processes and average differences in height between SCZ-cases and controls. This schematic overview only presents a plausible hypothesis for integrating the results of this study. TSH: thyroid stimulating hormone. Created with BioRender (*[*www.biorender.com*](http://www.biorender.com)*) with permission to publish.*

### **Supplementary Figures**

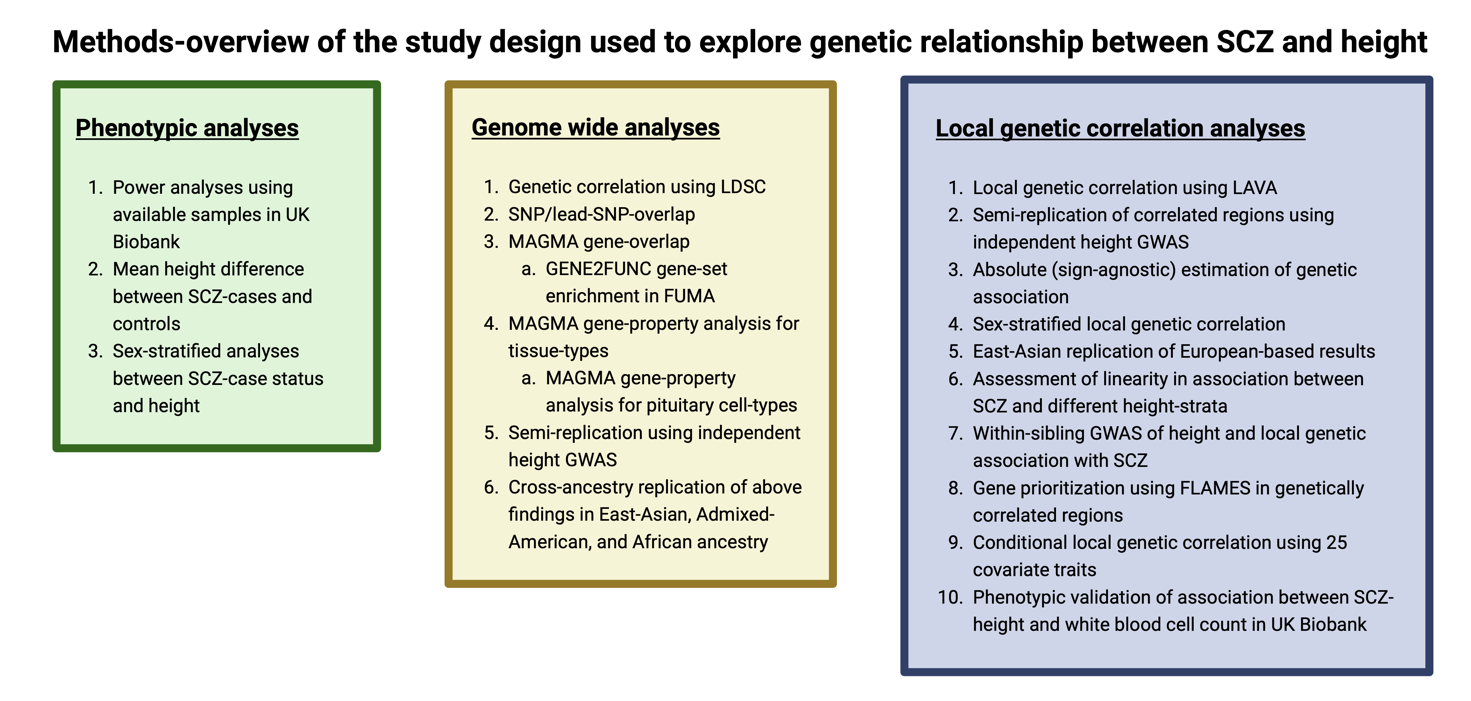

#### **Supplementary Figure 1. Overview of the study design**

*To explore the SCZ-height relationship, we conducted phenotypic, genome-wide and local genetic analyses. Phenotypic analyses were used to replicate a previously reported mean difference in height between SCZ cases and controls. Genome-wide analyses were conducted to find overlapping SNPs, genes, and gene-property results between SCZ-height, as well as to semi-replicate these findings in an independent height cohort (while using the same SCZ summary statistics), and to replicate findings in three different ancestry groups. Lastly, local genetic correlation analyses were conducted to explore both positive and negatively associated genomic loci between SCZ and height. Created with BioRender (*[*www.biorender.com*](http://www.biorender.com)*) with permission to publish.*

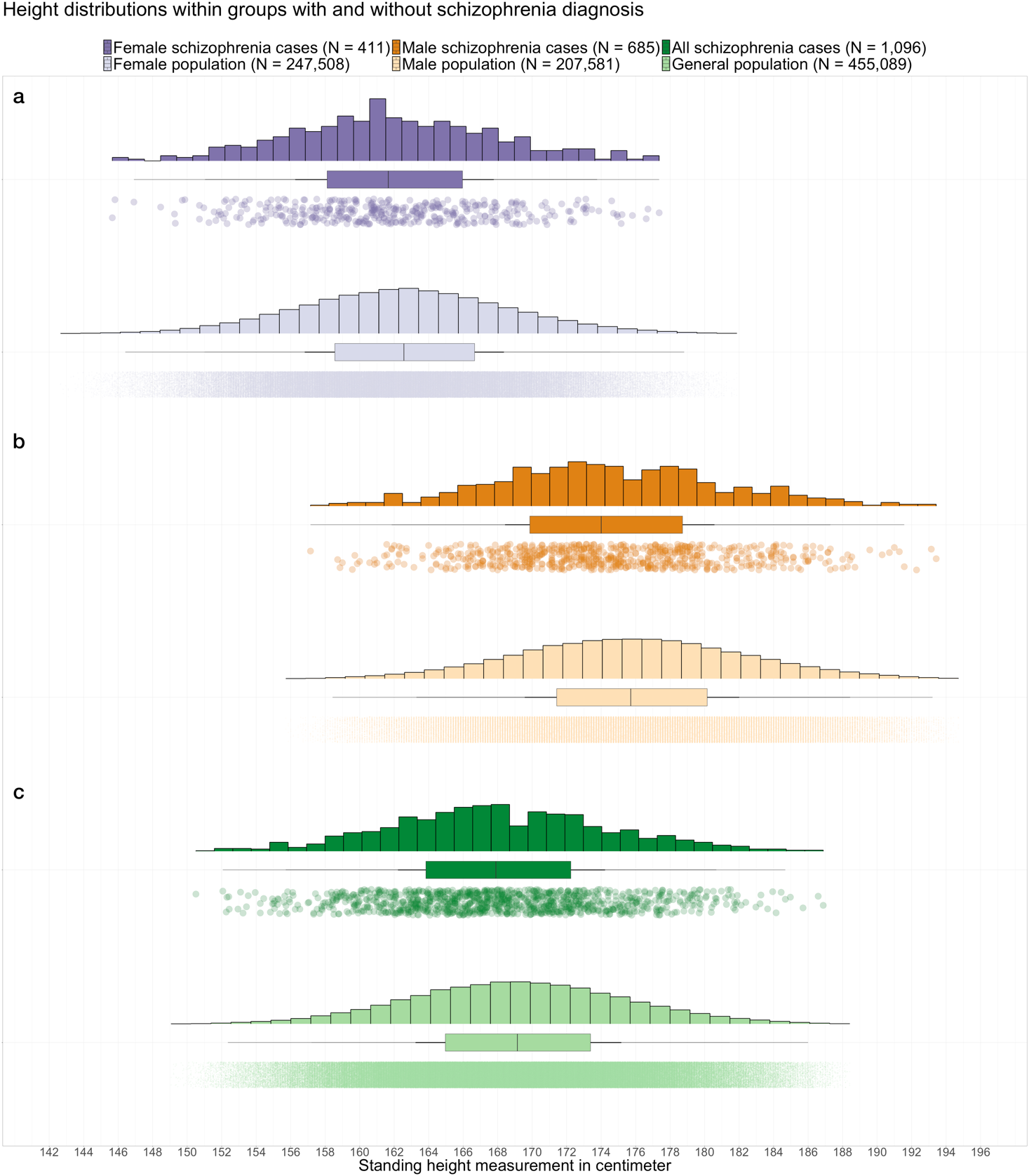

#### **Supplementary Figure 2. Distribution of height comparing subjects with and without a diagnosis for schizophrenia within females-only, males-only and all subjects**

*Distribution of age-adjusted height values for females (a, purple), males (b, orange) and combined (c, green) samples between subjects with (darker color) and without (lighter color) a SCZ diagnosis. Height data of the combined sample were both age- and sex-adjusted. Upper and lower hinges of the boxplot represent the 1^st^ and 3^rd^ quartiles (or interquartile range) and the upper and lower whiskers from the hinges are the interquartile range (i.e., ­±1.5 x IQR). The distribution of points underneath all boxplots represents the individual data points from each subject within their group. In all cases, the SCZ case group showed a slight shift to the left, indicating the 0.79 to 1.29 cm shorter mean height compared to controls.*

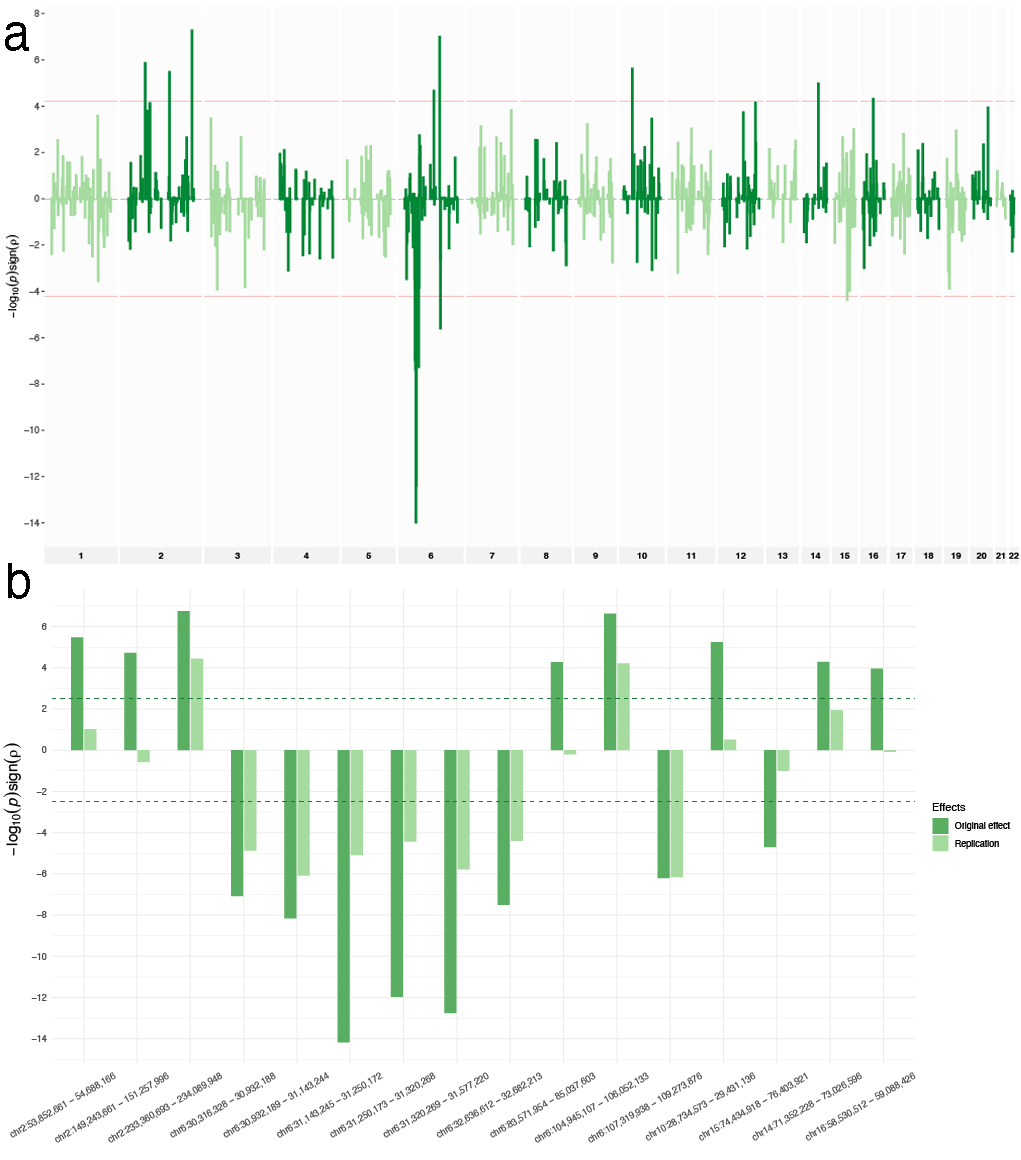

#### **Supplementary Figure 3. Local genetic correlations and replication between SCZ and height**

*Genetic correlations between SCZ and height,* *with genomic location on the x-axis and the -log10 of the p-values corresponding to the correlations on the y-axis (red dotted line indicating the genome-wide significance threshold for positive (top) or negative (bottom) correlation estimates, respectively). a) All 816 estimated SCZ-height genetic correlations across the genome (x-axis); b) The initial 16 significant genetic SCZ-height correlations (dark green) and corresponding genetic correlations observed in a semi-replication attempt using a different independent GWAS on height (light green). Out of the 16 correlations, 9 were replicated and further analyzed in downstream analyses.*

**Supplementary Figure 4. Locus zoom plot for regions with shared prioritized genes**

*SNPs are dots in locus zoom plot and coloured by their LD R^2^ value with reference to the top prioritized SNP in the locus. Plots are shown for the same locus for SCZ and height, and top prioritized SNPs for both traits are highlighted in each plot.*

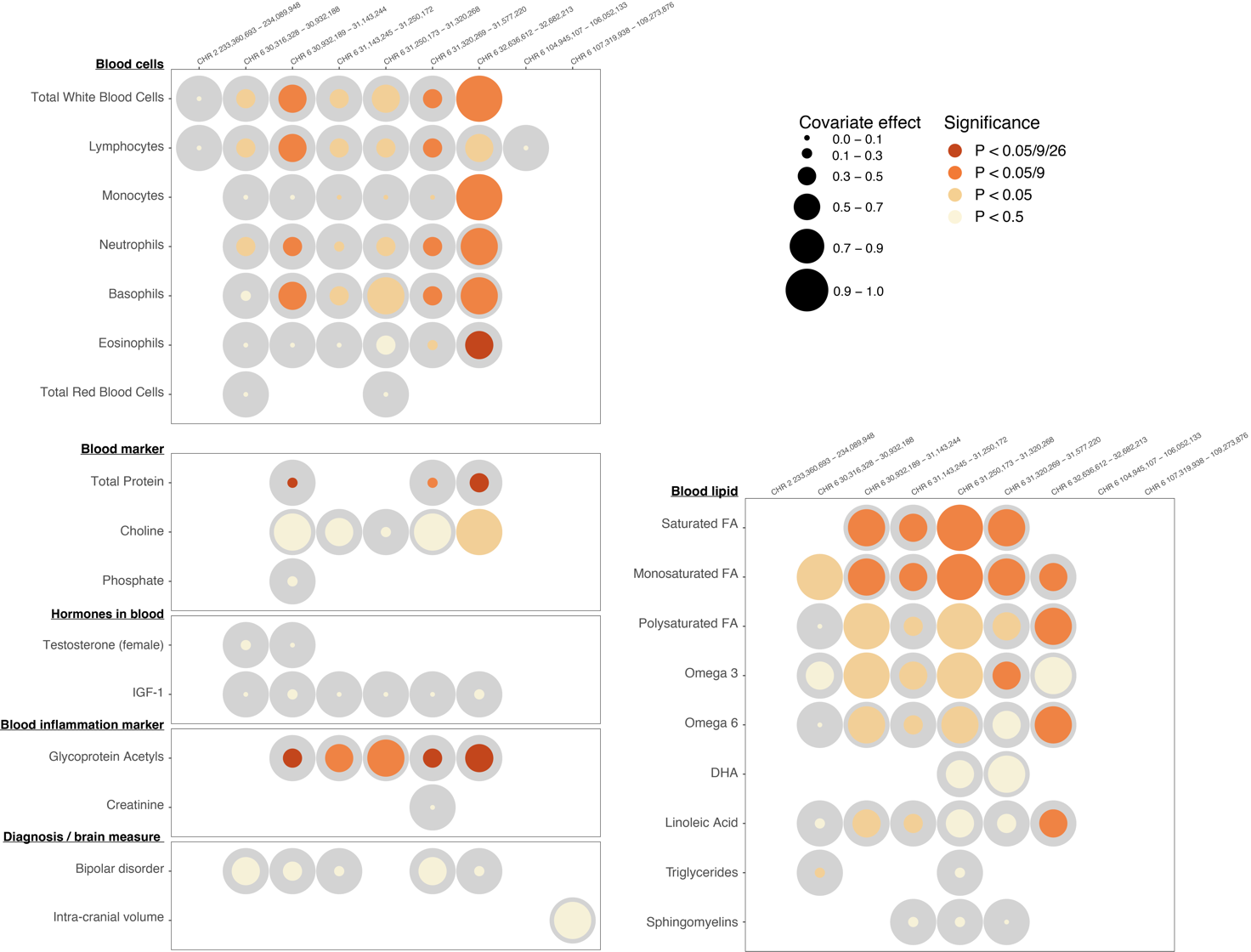

#### **Supplementary Figure 5. All results from conditional local genetic correlation analysis with height, rather than SCZ, as outcome**

*Results of the conditional genetic correlation analyses for all 25 covariates. Left side of the plot shows covariate name and category; top side gives genomic region definitions. Circle sizes represent the estimated covariate effect: the covariate effect is the estimated genetic covariance between the covariate and the covariance shared between SCZ-height, where grey circles sizes are a reference for what size represent complete overlap. Colored circles are the actual obtained genetic covariance between covariate and SCZ-height within each of the 9 genomic regions. The significance of the covariate is given by color, where light yellow = not significant, light orange = nominally significant, orange = significant corrected for number of genomic regions tested (9), and dark red = significant corrected for number of genomic regions and number of covariates tested (9 x 25).*

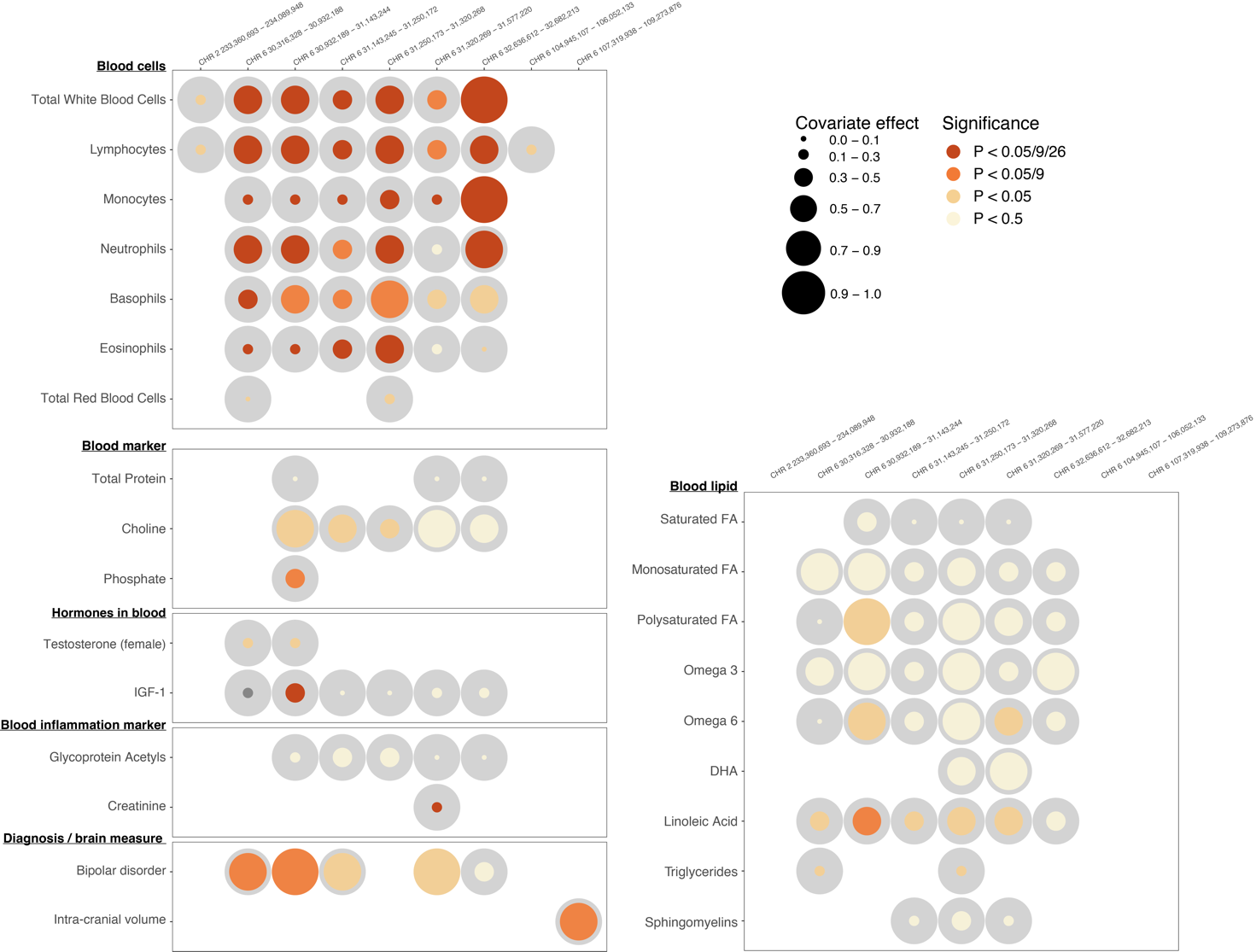

#### **Supplementary Figure 6. All results from conditional local genetic correlation analysis with SCZ as outcome**

*Results of the conditional genetic correlation analyses for all 25 covariates. Left side of the plot shows covariate name and category; top side gives genomic region definitions. Circle sizes represent the estimated covariate effect: the covariate effect is the estimated genetic covariance between the covariate and the covariance shared between SCZ-height, where grey circles sizes are a reference for what size represent complete overlap. Colored circles are the actual obtained genetic covariance between covariate and SCZ-height within each of the 9 genomic regions. The significance of the covariate is given by color, where light yellow = not significant, light orange = nominally significant, orange = significant corrected for number of genomic regions tested (9), and dark red = significant corrected for number of genomic regions and number of covariates tested (9 x 25).*

### **Supplementary material references**

19.	Schizophrenia and Inflammation Research: A Bibliometric Analysis - PMC. https://www.ncbi.nlm.nih.gov/pmc/articles/PMC9219580/.
